# Statistical Inference Using GLEaM Model with Spatial Heterogeneity and Correlation between Regions

**DOI:** 10.1101/2022.01.01.21268139

**Authors:** Yixuan Tan, Yuan Zhang, Xiuyuan Cheng, Xiao-Hua Zhou

**Affiliations:** Department of Mathematics, Duke University; School of Mathematical Sciences, Peking University; Center for Statistical Sciences, Peking University; Beijing International Center for Mathematical Research, Peking University; Department of Biostatistics, School of Public Health, Peking University

## Abstract

A better understanding of the various patterns in the coronavirus disease 2019 (COVID-19) spread in different parts of the world is crucial to its prevention and control. Motivated by the celebrated GLEaM model (Balcan et al., 2010[1]), this paper proposes a pioneering stochastic dynamic model to depict the evolution of COVID-19. The model allows spatial and temporal heterogeneity of transmission parameters and involves transportation between regions. Based on the proposed model, this paper also designs a two-step procedure for parameter inference, which utilizes the correlation between regions through a prior distribution that imposes graph Laplacian regularization on transmission parameters. Experiments on simulated data and real-world data in China and Europe indicate that the proposed model achieves higher accuracy in predicting the newly confirmed cases than baseline models.

## 1 Introduction

The outbreak of coronavirus disease 2019 (COVID-19) has impacted all aspects of the world significantly for a long. As of 26 Oct 2021, there have been over 243 million confirmed cases of COVID-19, including over 4 million deaths [2]. Therefore, it is essential to study the spread of COVID-19 for better prediction and prevention of the disease. This paper first proposes a stochastic dynamical model that describes the spread of COVID-19 in multiple regions and then develops an algorithm to estimate the transmission parameters and their posterior distributions.

The model proposed in this paper is inspired by The Global Epidemic and Mobility (GLEaM) model proposed in [1]. GLEaM is a stochastic dynamic model to depict the spread of epidemics, integrating multiple data layers that couple metapopulations at the global level. The model involves 3362 subpopulations in 220 countries obtained from Voronoi tessellation, centered around major airports. These subpopulations are connected by a multi-layer mobility network composed of processes from short-range communication in geographically close subpopulations to international flights. In each subpopulation, the transmission of epidemics is modeled by a Susceptible-Exposed-Infected-Removed (SEIR) compartmental model [3] that integrates short-range communication. After specifying the compartmental model and initial conditions, GLEaM can be run repeatedly to build a statistical ensemble for each point in the space of parameters, which can be further utilized for Monte Carlo analysis of epidemic parameters. A more detailed review of SEIR model and the GLEaM model is in Appendix A.

In the vast majority of GLEaM’s applications [4, 5, 6, 7, 8, 9, 10, 11], the parameters are estimated based on [12]. [12] performed the maximal likelihood analysis of the reproduction number *R*_0_ in the seed region Mexico. For each value of the reproduction number *R*_0_, the method generated the distribution of arrival time of the influenza A(H1N1) in 12 countries produced by 2 × 10^3^ GLEaM simulations. Then, the optimal the reproduction number *R*_0_ was chosen by maximizing the likelihood function of arrival time. [12] and subsequent works following its settings [4, 5, 6, 7, 8, 9, 10, 11] assumed that the epidemic was seeded from one region and the transmission parameters or the other parameters (like the introduction date and location) were estimated through the maximal likelihood analysis of arrival time or other events. In particular, the method in [12] was adopted in [6] to estimate the posterior distribution of the reproduction number *R*_0_ of COVID-19, which was assumed to be uniform for all subpopulations at all times. However, this setting is not suitable for the current scenario of the COVID-19 pandemic, since COVID-19 has lasted for a long time, and the community transmission has been widespread in most countries in the world [13]. To model the spread of COVID-19, both spatial and temporal heterogeneity of the transmission parameters are needed instead of directly modeling the reproduction number *R*_0_ as a periodic function of time as in [12], since the social behaviors, containment measures, medical conditions, and other elements that might have effects on the spread of COVID-19 may vary among different countries and over time.

Recently, [14] improved the model described above by estimating the initially infected individuals in each subpopulation through microblogging data from Twitter and also estimating the reproduction number *R*_0_ for USA, Italy, and Spain separately. Therefore, the method used in [14] involved spatial heterogeneity. However, in this study, international travel was not considered, and GLEaM was applied to each of the aforementioned countries (as an isolated system) independently. The transmission rates in all subpopulations of these countries were again presumed to be homogeneous. Furthermore, [14] also assumed that the initially infected individuals for each subpopulation were proportional to the total number of Twitter users in that subpopulation. Thus it still assumed that the severity of the pandemic at the initial outbreak of COVID-19 was uniform over the country, which is not the case for COVID-19.

In addition to the issues mentioned above, other potential concerns exist when using GLEaM to model the spread of COVID-19. As mentioned in the last section of [15], GLEaM can be used to simulate the spread of the epidemic under normal conditions since it uses the “steady-state” mobility data around the world. However, since the outbreak of COVID-19, the social order has been disrupted, and travel has been restricted in most countries. Thus GLEaM might not work well with its multi-layer mobility networks. Furthermore, the estimate of parameters using GLEaM is based on a large number of simulations to explore the space of parameters, which usually takes much computational time [1]. In addition, although the social behavior, medical conditions, and other factors that affect the spread of COVID-19 may vary among different regions, these factors for regions that are geographically close or have similarities in other aspects still bear some resemblance. Hence, the transmission rates for COVID-19 should not only have their own heterogeneity but also correlate with each other. However, neither of the features is reflected in GLEaM or most of its applications.

As the consequences of the possible constraints of GLEaM described above, most of the papers using GLEaM to model the epidemics mainly focus on estimating only the transmission parameter in the “seed region” at the very beginning of the outbreak. However, for the current long-lasting spread of COVID-19 pandemic all over the globe, the spatial and temporal heterogeneity of the transmission parameters is needed to be taken into full consideration.

In this paper, we propose a pioneering stochastic model based on the GLEaM framework that enables transmission parameters’ spatial and temporal heterogeneity. In contrast to most applications of GLEaM, which mainly focus on the initial outbreak, the model we propose is able to model the long-lasting spread of COVID-19. For the inference of the parameters, this paper introduces an optimization algorithm that utilizes the correlation between districts. Furthermore, the posterior distribution of parameters is estimated by Markov chain Monte Carlo (MCMC) sampling starting from an optimal choice of the parameters obtained from the optimization process. This takes a much shorter time than the methods based on GLEaM simulations.

The main contributions of our paper are:

- We propose a new stochastic model to describe the epidemic’s lasting spread, allowing spatial and temporal heterogeneity of transmission parameters and transportation between districts.
- Based on the model, we also design an algorithm that first makes inference for the parameters through a two-step procedure combining the information of correlation between districts, which is equivalent to imposing graph Laplacian regularization on the transmission parameters. Then, the algorithm estimates the posterior distribution efficiently by MCMC sampling with the estimated parameters as the initial points.
- We compare the performance of the proposed model with the baseline models on both simulated data and real-world data.
  - For the simulated data, the results show that combining heterogeneity and transportation into the model helps improve the performance of trajectory prediction and parameter estimation. Moreover, our inference algorithm that integrates the correlation of districts leads to further improvement in predicting the future trajectories.
  - For the real-world data in China and Europe, the proposed model still outperforms the baselines in trajectory prediction.

Data sets used in this paper are publicly available. Our work focuses on proposing a new method for parameter inference and, consequently, answering questions regarding the long-lasting spread of COVID-19 in multiple regions.

A strength of the proposed model resides in introducing spatial and temporal heterogeneity of transmission parameters. We remark that there exist recent works which also involved different levels of heterogeneity in their models in various ways. [16, 17] utilized randomness in reproduction number to reflect heterogeneity of the population, using plate model with Bayesian method and heterogeneous well-mixed theory [18] with age-of-infection method [19], respectively. [20] and [21] used functional data analysis tools. Specifically, [20] captured two different epidemic patterns in different regions of Italy using probKMA algorithm [22], and [21] revealed different patterns of the epidemic across countries with functional principle component analysis. In addition, [23, 24, 25, 26, 27] adapted SEIR / Susceptible-Exposed-Infected (SEI) / Susceptible-Infected (SI) compartmental models similar to this paper. Among these works, [23, 24, 25] considered heterogeneity in the aspects of age groups, social links, and vaccination status separately. [26, 27], including spatial heterogeneity in one county and among states respectively, bore more similarity with this paper since they also allowed transmission parameters to be spatially heterogeneous and involved transportation between different regions. However, [26] only considered intracounty data, and the transportation was used to compute the effective size of compartments and did not affect the dynamic model. Furthermore, the transmission rates in [26] were determined by an SDE whose parameters were to be fitted. Therefore, [26] focused on a different scope from this paper. The settings of compartments in [27] are more realistic than the one considered in this paper by considering reporting rates. Nevertheless, compared with the model and inference algorithm described in Section 2, transmission rates in [27] were kept constant over time and were estimated using independent non-informative prior. Both [26] and [27] used Ensemble Kalman Filter, which samples particles in the state space according to the prior distribution and obtains the posterior distribution in the process of moving particles at each time step. This may be computationally less efficient than directly applying MCMC according to the posterior distribution with the initial point maximizing the likelihood function, as implemented in this paper.

We list the default notations and parameters used throughout the paper in Table 1. The rest of the paper is structured in the following way: In Section 2, we introduce the stochastic dynamic model and corresponding inference algorithm. In Section 3, we compare the performance of trajectory prediction and parameter estimation of models with or without mobility, heterogeneity, or using correlation information in the inference part for the simulated data. Section 4 describes the real-world data used in this paper, presents the results and findings of applying our model to the COVID-19 data in China and Europe. We discuss our limitations and possible extensions in Section 5.

**Table 1:**
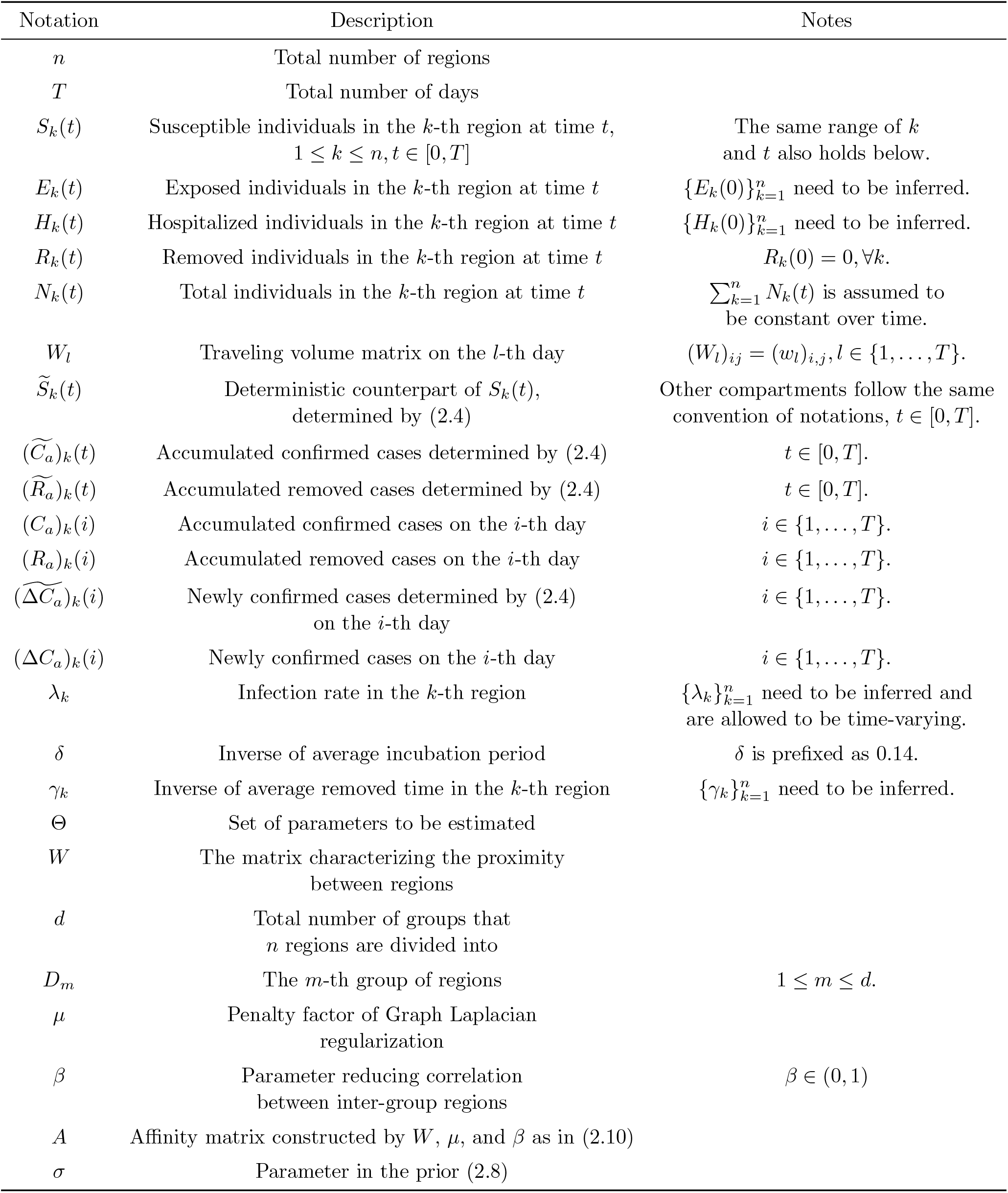
List of notations and parameters

## 2 Method

### 2.1 Ethics Statement

The medical record data in China and Europe used in this paper are publicly available and from the official websites of the National Health Commission of the People’s Republic of China [28], Chinese Center for Disease Control and Prevention [29], and European Centre for Disease Prevention and Control [30]. The collection of data is performed in compliance with local government regulations. More details of data sources can be found in Section 4.1.

### 2.2 Model Description

#### Compartmental model over multiple regions

In GLEaM ([1]) and other works ([26, 27]) involving transportation, the whole area is first divided into subdivisions. For example, the GLEaM model divides the total area of 220 countries into over 3300 subpopulations centered around major airports and [26] divided Milwaukee County and Dane County in the state of Wisconsin into several regions. In this paper, we consider abstract subdivisions in the whole area, which will be referred as “regions” hereinafter until further specifications in the later experiment sections. We denote by *n* the number of regions.

In our model, we use continuous time *t* ∈ [0, *T* ], where it is assumed that the evolution of the epidemic lasts within a period of *T* time units. The unit of time is fixed as one day throughout this paper. Note that when we introduce the transportation model in below the traveling matrix is assumed to be constant within each day, and also the observed data is collected on a daily basis. Thus we will use notation of discrete time (days) from 1, …, *T* hereinafter, however, the evolution dynamic itself is modeled over continuous time.

For each region, we consider the following epidemiology compartments adapted from the SEIR model:

- *S*_*k*_(*t*): Susceptible.
- *E*_*k*_(*t*): Exposed and infectious.
- *H*_*k*_(*t*): Hospitalized.
- *R*_*k*_(*t*): Removed.

The subscript *k* of the states denotes that they belong to the *k*-th region, and dependence on the continuous time *t* is addressed through expressing the states as functions of *t* ∈ [0, *T* ]. Removed individuals are those who have recovered or died.

At time *t*, we use *N*_*k*_(*t*) = *S*_*k*_(*t*)+*E*_*k*_(*t*)+*H*_*k*_(*t*)+*R*_*k*_(*t*) to denote the total population in the *k*-th region. *N*_*k*_(*t*) is allowed to be time-varying since the inter-region mobility is taken into consideration, especially for the days before the implementation of travel restrictions. However, since the traveling volume is not comparable to the total population in a region, the fluctuation of the total population in a region is not obvious. In this paper, we assume that 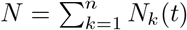 keeps constant over time, which means that we consider a closed system in this paper, where exported/imported cases are not considered. However, it is worth noting that we do allow the transportation of active virus carriers between regions within our system. We remark in advance that this assumption is reasonable for the real-world data sets considered in this paper. From January to February 2020, strict international travel restrictions were imposed in China. While for data in Europe, from May to August 2020, the local spread of the epidemic has reached a relatively high level, and the imported cases were not comparable to the indigenous cases. We also denote (*N*_*a*_)_*k*_(*t*) = *S*_*k*_(*t*) + *E*_*k*_(*t*) + *R*_*k*_(*t*) as the total population that are permitted to move in the *k*-th region, excluding the hospitalized ones.

#### Transportation between regions

Transportation plays an essential role in the spread of COVID-Actually, [31, 32] indicated that the travel restrictions were remarkably important in mitigating the transmission of COVID-19, especially in the early stage of the pandemic. Recently, as detailed in [33], the Omicron variant had spread to 110 countries and had become dominant in many of them by 22 December 2021, only one month after its first report from South Africa on 24 November 2021. This motivates us to also take transportation into consideration in this paper. Motivated by the GLEaM model [1], we introduce the transportation between regions. Specifically, we denote (*w*_*l*_)_*ij*_ as the traveling volume from region *i* to *j* on the *l*-th day (*l* = 1, …, *T*). Then the traveling matrix *W*_*l*_ on the *l*-th day can be written as

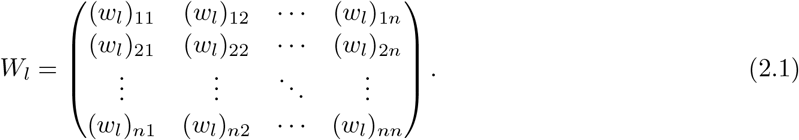

Then, inspired by [34], we consider the transition dynamic of *ξ*(*t*) = {*S*_1_(*t*), *E*_1_(*t*), *H*_1_(*t*), *R*_1_(*t*), …, *S*_*n*_(*t*), *E*_*n*_(*t*), *H*_*n*_(*t*), *R*_*n*_(*t*)} ∈ {0, 1, 2, …, *N* }^4*n*^ over *t* ∈ [0, *T* ] described below:

- Transmission in the *k*-th region: A case from *E*_*k*_(*t*) chooses an individual from *N*_*k*_(*t*) randomly at Poisson rate *λ*_*k*_(*t*), and the individual chosen is infected if it is of state *S*_*k*_(*t*). Note that this is different from traditional SEIR models, since we assume that for the COVID-19 case, the pre-symptomatic patients from *E*_*k*_(*t*) can be contagious.
- Hospitalization in the *k*-th region: Each individual in *E*_*k*_(*t*) will be hospitalized with Poisson rate *δ*.
- Recovery or death in the *k*-th region: Each individual in *H*_*k*_(*t*) will transfer into *R*_*k*_(*t*) with Poisson rate *γ*_*k*_. The rate *γ*_*k*_ owns spatial heterogeneity due to the uneven distribution of medical resources.
- Transportation between regions: At the end of the *l*-th day, all the individuals in region *i* except the ones in *H*_*i*_(*l*) have the same probability to travel from the *i* region to the *j* region, and the total traveling volume from the *i* region to the *j* region is (*w*_*l*_)_*ij*_. We assume that there are no transmissions happening during the transportation between regions. If we denote 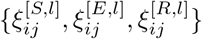 as the number of people transported from {*S*_*i*_(*l*), *E*_*i*_(*l*), *R*_*i*_(*l*)} to {*S*_*j*_(*l*), *E*_*j*_(*l*), *R*_*j*_(*l*)} at the end of the *l*-th day, then 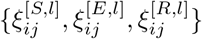 follows a multinomial distribution. Specifically,

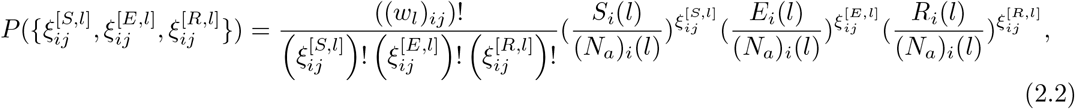

with 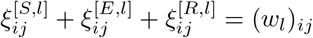.

As a consequence, *N*_*k*_(*t*) is a piece-wise constant function of *t* which only changes at the end of each day. Specifically, for any time *t* ≥ 0,

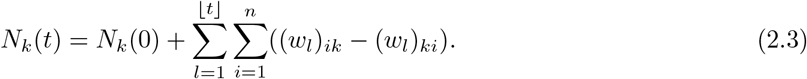

#### Differential equation with spatial heterogeneity

Following [35, 36], we consider the mean-field differential equation system defined by (2.4), which is a deterministic version of the stochastic model above:

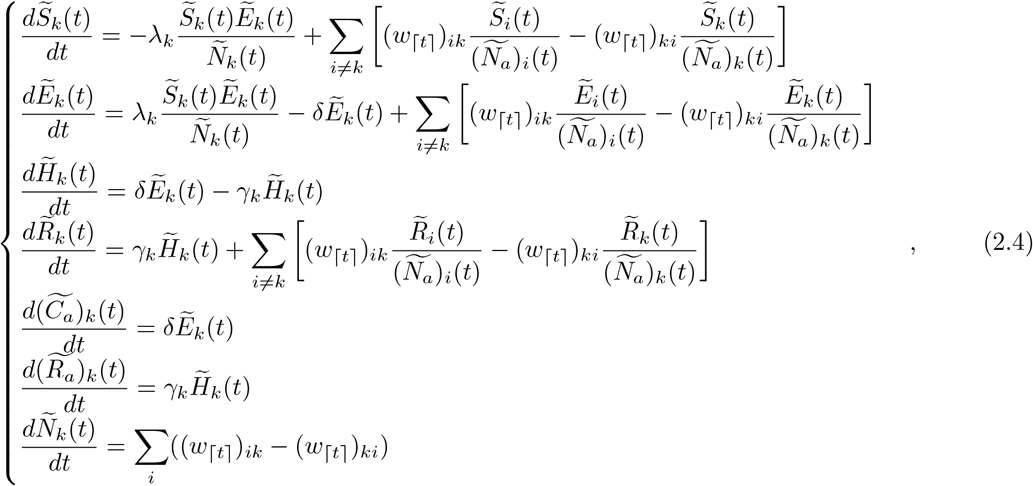

where 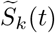 is the deterministic counterpart of *S*_*k*_(*t*) and so on. Note that since the traveling volume is a piece-wise constant function of *t*, 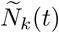 evolves as a piece-wise linear function of *t*, and 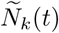 coincides with *N*_*k*_(*t*) when *t* takes integer values. 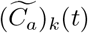 and 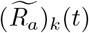 are the accumulated confirmed and removed cases determined by the ordinary differential equation (ODE) system (2.4) in the *k*-th region at time *t* respectively. 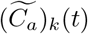 (or 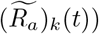 only takes the increment of 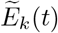 (or 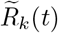 into account, meanwhile guaranteeing that each case is only accounted for once.

Furthermore, we assume that the accumulated confirmed and removed cases are available from data on a daily basis, which are denoted as 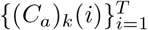 and 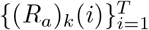, respectively. We also assume that *R*_*k*_(0) is 0, while *E*_*k*_(0) and *H*_*k*_(0) are left to be inferred for each *k* = 1, …, *n*. For inference of parameters, we further denote 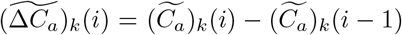 as the newly confirmed cases on the *i*-th day (*k* = 1, …, *n, i* = 2, …, *T*) determined by (2.4), and (Δ*C*_*a*_)_*k*_(*i*) as the newly confirmed cases computed from data, namely (Δ*C*_*a*_)_*k*_(*i*) = (*C*_*a*_)_*k*_(*i*) − (*C*_*a*_)_*k*_(*i* − 1). The same convention holds for the definitions of 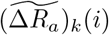 and (Δ*R*_*a*_)_*k*_(*i*). Note that the data 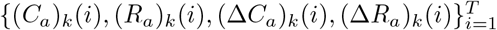are random in nature.

Note that the model and the inference algorithm described below can be applied to estimate spatially heterogeneous transmission parameters as long as 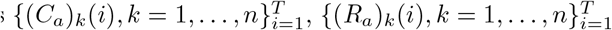, and 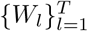 are available. The availability of 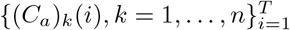 and 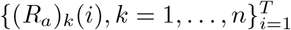 is required in many works that use the SEIR model to estimate transmission rates of the epidemic [37, 38, 39], and the transportation network is also used in GLEaM [1] and its applications. However, in contrast to the works based on GLEaM [12, 4, 5, 6, 15], we allow parameters to possess both spatial and temporal heterogeneity, and further utilize the correlation between regions in the inference of parameters. We remark that the spatial and temporal heterogeneity is reflected in the fact that the transmission parameters {*λ*_*k*_} are allowed to vary in both space and time in our model. The temporal heterogeneity is introduced in more detail for the real-world data in Section 4.2.

### 2.3 Estimation of Model Parameters

A two-step procedure of estimating parameters is described below.

First, note that *δ*, the inverse of the average time for a person from being exposed to hospitalized, is prefixed to be *δ* = 0.14 for all regions and all time. According to [40], the mean duration of incubation period is 5.2 days. Furthermore, we assume that the average time for an individual from showing symptoms to being hospitalized is 2 days for simplicity. Thus, the mean duration for an individual from being exposed to being hospitalized is 7.2 days, whose inverse value is approximately 0.14.

Thus, the model parameters that need to be estimated are 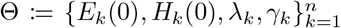. We also denote 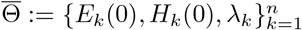, which excludes 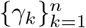 from Θ.

A two-step analysis is adapted for the estimation. We first make inference for 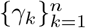 utilizing the newly removed cases and the accumulated hospitalized cases. After the estimation of 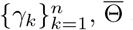 are then estimated by maximizing the posterior distribution, where we introduce a prior distribution combining the information of correlation between regions. In addition, the marginal posterior distributions of 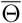 can also be obtained by running MCMC sampling starting from the optimizer 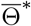of the posterior distribution. Then we complete the inference of 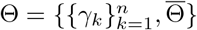.

Mathematically, given parameters Θ, we can run the ODE system (2.4) on [0, *T* ] and denote 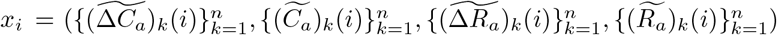 as the values determined by (2.4) on the *i*-th day, and 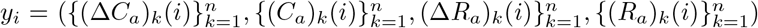 as the observed coun-terpart of *x*_*i*_.

#### Step 1: Estimate 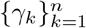

We first estimate 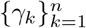 by maximizing 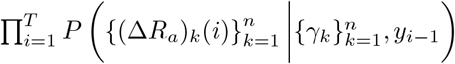over *γ*_*k*_ for each *k*.

By assuming that the newly removed cases in one day follow a Poisson distribution whose mean equals to the product of *γ*_*k*_ and the accumulated hospitalized cases (which is the difference between the accumulated confirmed cases and the accumulated removed cases, and thus is observable) the day before, we have that

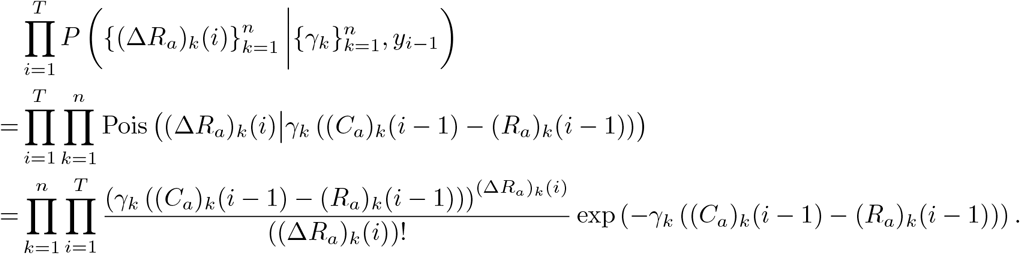

Then, we estimate 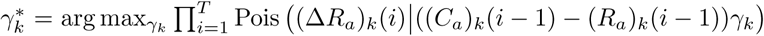 for each *k* sep-arately.

#### Step 2: Estimate 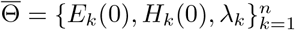 and the marginal posterior distributions of 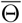

Next, we estimate the remaining parameters 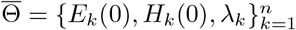 as follows.

First, note that *λ*_*k*_ is the transmission rate, which reflects the infectivity of the virus, the prevention and control measures, and the rates of close contacts in the *k*-th region.

Notice that the ODE system (2.4) is the mean-field version of our stochastic model, and 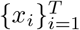 (especially 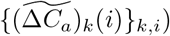 are determined by the parameters, thus by the Markov property, {(Δ*C*_*a*_)_*k*_(*i*)} are all independent for *k* = 1, …, *n, i* = 1, …, *T* conditioned on the parameters 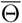. Furthermore we suppose that 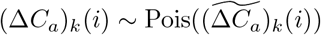. Thus, the likelihood of 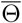 can be written as

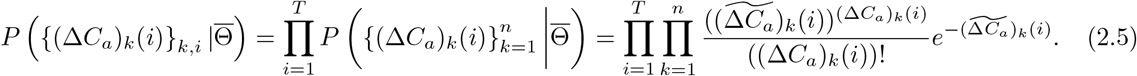

Now, we denote the posterior distribution of 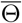 as 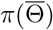. Then by Bayesian formula,

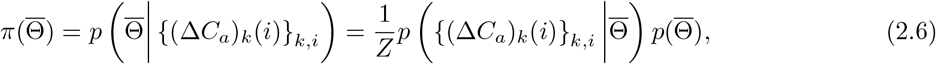

where 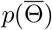 is the prior distribution of 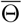 to be determined and *Z* = *p* {(Δ*C*_*a*_)_*k*_(*i*)}_*k,i*_ is a constant irrelevant to 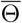. We further denote 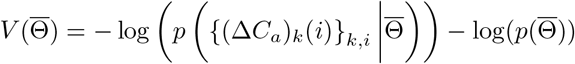, then

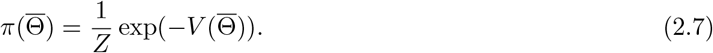

To fit the realistic evolution of the epidemic more precisely, 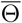 is estimated as 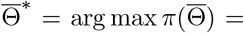 arg min 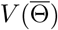 with reasonable prior distribution 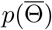. Then, MCMC sampling scheme starting from 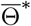 is applied to get the posterior distribution for 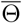. This process might possess higher computational efficiency than choosing the initial point for MCMC randomly or empirically.

##### Choosing prior distribution

The remaining problem is to choose the prior distribution 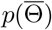. Presuming that the transmission rates in the regions owning more similarities are closer, 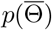 is designed to combine the information of correlations between regions. In particular, given a matrix *A* which characterizes the pairwise similarities between the regions, and if we denote *λ* = (*λ*_1_, …, *λ*_*n*_)^*T*^ ∈ ℝ^*n*^,

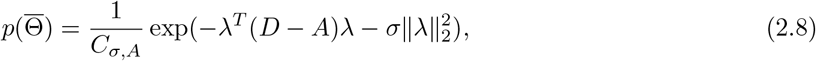

where *D* = diag{*d*_1_, …, *d*_*n*_} is the degree matrix of *A* with 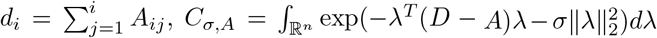 is a constant depending on *σ* and *A*. Here, a small *σ* is chosen for 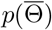 to be a probability measure without imposing much restriction on *λ*. Then,

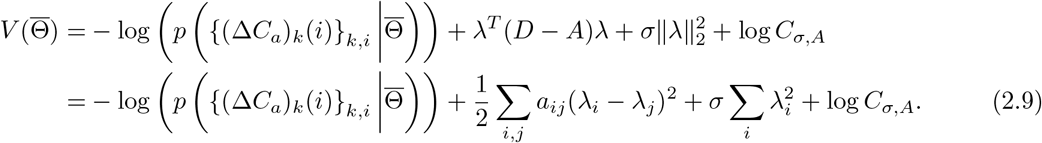

The parameter estimation procedure could be extended to the case when {*λ*_*k*_} are time-varying by modifying (2.11), which we will introduce in more detail in Section 4.2 for the real-world data in China and Europe.

*A* is constructed from affinity matrix *W* by further addressing the correlation between regions with more similarities. To treat different data sets and *W* with a unified approach, we assume that max_*i,j*_ *W*_*ij*_ = 1 (*W* can be re-scaled entry-wise if necessary).

For a given *W* = (*W*_*ij*_)_*i,j*∈{1,…,*n*}_ whose choice is detailed later, the next step of attaining *A* is to divide the *n* regions into *d* groups (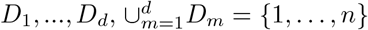, and ∀*i* ≠ *j, D*_*i*_ ∩ *D*_*j*_ = ∅) where the regions in the same groups have more similarities. Then, for given *β* ∈ (0, 1) and a given penalty factor *µ >* 0, *a*_*ij*_ is constructed as follows:

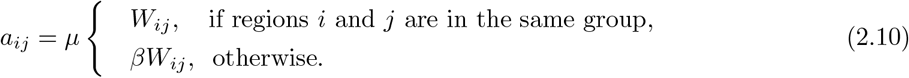

We remark that *β* in (2.10) is taken to be 0.1 for all the experiments in this paper. By constructing *A* as in (2.10), correlations for regions in the same groups are further strengthened, whose transmission parameters are imposed with stronger restrictions.

Now we specify the choice of *W* for data sets that will be analyzed later in this paper. For simulated data and real-world data in China, in which cases the transportation data are available, we construct *W* from the traveling volume matrices 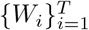. Specifically, 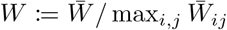, where 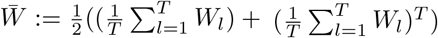. Nevertheless, for real-world data in Europe, where we are not aware of traveling data publicly available that are sufficient for the proposed model, *W* is just the all 1 adjacency matrix. We remark that the affinity matrix *W* may also be obtained by ways other than using the transportation data, as long as it reflects the similarities between districts.

From the definition of *a*_*ij*_ in (2.10), the inference of 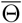 can be equivalently written as follows

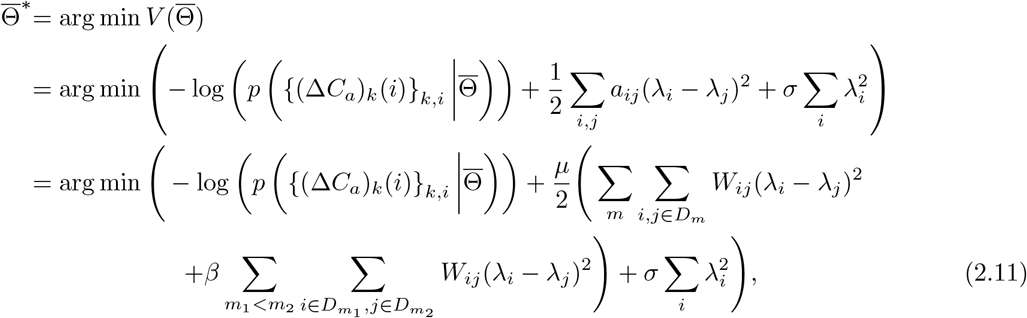

it can be seen that by choosing the prior distribution as in (2.8), a *l*_2_ regularization term 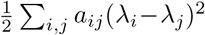 is imposed for better generalization.

Finally, after choosing 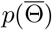 determined by *A* and *σ*, the optimization process 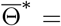 arg min 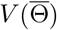 is accomplished through Matlab function `fmincon`. In addition, the posterior distribution for 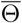 could be obtained by classical MCMC sampling scheme starting from 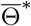 as mentioned above.

We summarize the procedure described in Section 2.3 in Algorithm 1 below.

###### Algorithm 1 GLEaM model with Graph Laplacian Prior Distribution

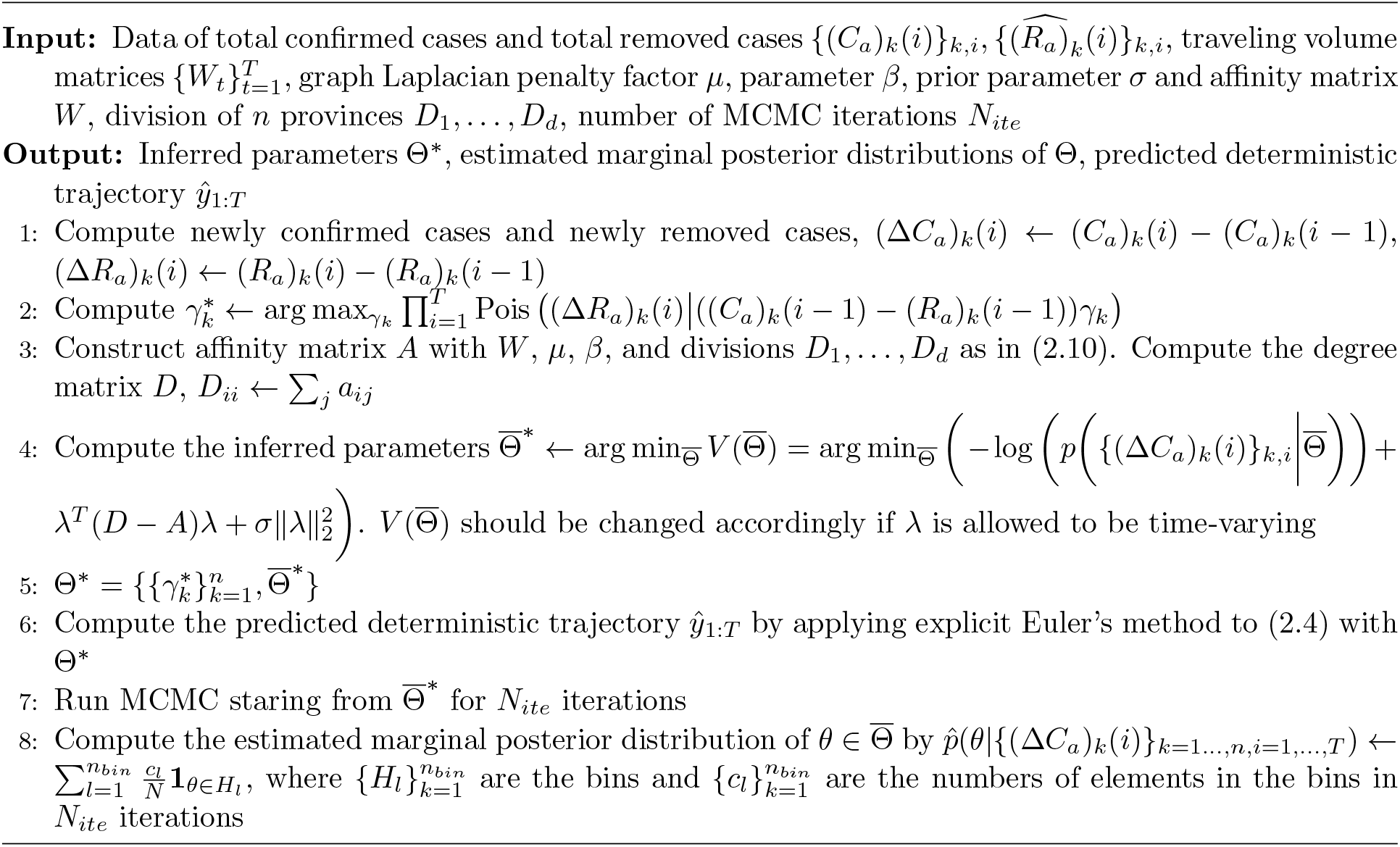

### 2.4 Prediction of the Epidemic Trajectories with the Estimated Parameters

Once the parameters 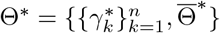 are estimated from the optimizations 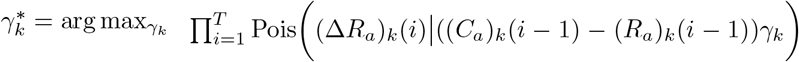 and 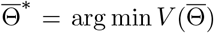 as described in Section 2.3, the trajectories of newly confirmed cases could be simulated according to the stochastic dynamic process with Θ^∗^. Furthermore, trajectories could also be sampled from the posterior distribution of Θ instead of using Θ^∗^ alone, which also takes the randomness from 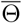 into account. Particularly, this could be achieved by sampling 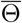 from MCMC and then simulating trajectories with the sampled 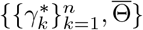.Additionally, deterministic trajectories determined by (2.4) could also be computed by explicit Euler’s method as in Algorithm 1.

## 3 Experimental Results for Simulated Data

Two specific cases are considered for simulated data. We first remark that the regions in Section 2 are called as provinces in this section. Section 3.2 considers four provinces separated into two groups (the provinces in the same group are assumed to have more similarities) and with traffic between each pair of the provinces. Section 3.3 considers thirty provinces randomly separated into three groups, with the other settings similar to the previous case. Section 3.1 includes more details of the experimental settings and sensitivity analysis.

### 3.1 More Details of Experimental Settings and Sensitivity Analysis

#### Experimental settings

The results in Section 3 are for 100 replicas. In each replica, three random trajectories are sampled independently according to the stochastic model with prefixed parameters, part of which are treated as the ground truth training, validation, and testing trajectory, respectively (see more details in Appendix B.1.1). For each model, we first fit the parameters using the training trajectory and then predict the testing trajectory using the estimated parameters. Note that we detail the choice of hyper-parameters for the model proposed in Appendix B.2. In particular, the penalty factor *µ >* 0 is chosen by cross-validation and chosen as the value minimizing the validation error, since for the simulated data, the validation error are identically distributed as the testing error. The comparison of trajectory prediction is from one typical realization, for which we compare the ground truth training and testing trajectories with the fitted training and predicted testing trajectories for all the models. Additionally, parameter estimation and quantitative evaluations are compared with mean and standard deviation over all 100 replicas. The detailed computations of training, validation, and testing errors can be found in Appendix C.

#### Sensitivity analysis of *σ*

Note that parameter inference with the proposed model involves the parameter *σ*, as shown in (2.11). Therefore, the sensitivity analysis for the parameter *σ* in (2.11) is performed for the four provinces case. Specifically, the results for *σ* varying from 10^−6^ to 10^0^ are presented and compared. Details can be found in Sections 3.2.4 and 3.2.5. Similar results are obtained for other data sets, and details are omitted.

#### Mismatched partition of regions

We also note that the graph Laplacian penalty of the proposed model depends on the partitioning the regions into several groups, as described in Section 2.3. Since the graph knowledge is usually not fully known, it is a question whether our methods can still perform well without accurate prior knowledge. For the thirty provinces case, we report the results of the proposed model with a mismatch between the partition of the regions and the ground truth division, the details of which can be found in Section 3.3.

### 3.2 Four Provinces Case

#### 3.2.1 Data Description

In this simulated study, we let *n* = 4, *T* = 20, and set the threshold *T*_*th*_ separating training and testing data to be 10 (more detailed can be seen in Appendix B.1.1). The other prefixed parameters are listed below:

- For *k* ∈ {1, 2, 3, 4}, *N*_*k*_(0) = 10^6^, *E*_*k*_(0) = 30, *H*_*k*_(0) = 10.
- For *l* ∈ {1, …, *T* }, *i, j* ∈ {1, 2, 3, 4} and *i* ≠*j*, (*W*_*l*_)_*ij*_ = 5 × 10^3^.
- *λ*_1_ = 0.5, *λ*_2_ = 0.47, *λ*_3_ = 0.4, *λ*_4_ = 0.37, *δ* = *γ*_1_ = … = *γ*_4_ = 0.14.

The four provinces are divided into two groups, with the first group consisting of province 1 and 2 and the second group consisting of province 3 and 4. The similarities within groups are reflected in the settings that the values of {*λ*_*k*_} are closer for provinces in the same group.

#### 3.2.2 Models to Compare

We first specify the models to be compared below. The last one is the proposed model, and the first four models serve as baselines with different settings.

1. The model with uniform prior distribution, without heterogeneity or migration.
2. The model with uniform prior distribution, without heterogeneity but with migration.
3. The model with uniform prior distribution, with heterogeneity but without migration.
4. The model with uniform prior distribution, with both heterogeneity and migration.
5. The model with prior distribution based on graph Laplacian, with both heterogeneity and migration.

For better illustration and comparison between the models in the experiment results, the Models 1-5 are summarized in Table 2 below.

**Table 2:** Models to be compared when the transportation data are available. Model 5 is the proposed model in this paper, and Models 1-4 are baseline models with different settings.

| Model | Migration | Heterogeneity | Prior of $\Theta$ |
| --- | --- | --- | --- |
| 1 | × | × | uniform prior |
| 2 | ✓ | × | uniform prior |
| 3 | × | ✓ | uniform prior |
| 4 | ✓ | ✓ | uniform prior |
| 5 | ✓ | ✓ | graph Laplacian prior |

First, the models with uniform prior distributions themselves (Models 1-4) are compared according to whether two key assumptions exist in the model:

1. Whether the transmission rates {*λ*_*k*_} are allowed to vary over regions.
2. Whether there exists transportation between regions.

Then, the model with prior distribution based on graph Laplacian (Model 5) is compared with those using uniform distributions as prior distributions (Models 1-4). The former one utilizes the correlation between subpopulations by adding a *l*_2_ regularization term for the model. In contrast, only lower and upper bounds are imposed on parameters without other prior information being used in the latter ones.

For Model 5, we conduct the sensitivity analysis for parameter *σ* in (2.8) and present results for *σ*= 10^−6^, 10^−3^ and 10^0^ respectively. Then for Model 5, by (2.11), 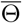 are estimated by the optimization problem in (3.1):

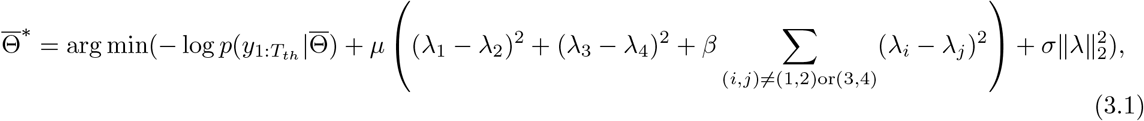

where *β* is taken to be 0.1.

We remark that in Models 1-4, the estimation of 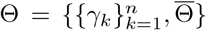 still follows a similar two-step procedure as in Model 5, and the first step of obtaining 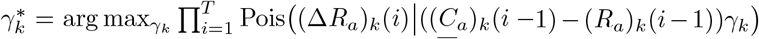 remains formally the same. The difference lies in the optimization object of 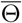. First, the *l*_2_ regularization term becomes prior knowledge of the parameters’ upper and lower bounds. Second, for models without heterogeneity of parameters, 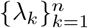 are forced to be the same in (2.4). For models without transportation between regions, terms involving *W*_*t*_ disappear in (2.4). Additionally, for the other data sets considered in the following sections, the parameter estimation methods for the baseline models are similar and thus will not be repeated.

Finally, note that the model in [12] is similar to Models 1 and 2, since they all assume a spatially homogeneous transmission parameter. However, [12] assumed that the epidemic was seeded from one seed region while Models 1 and 2 do not make such assumption. Moreover, [12] focused more on the spread of the epidemic from the seed region at the early stage of the pandemic, and only the introduction dates in the other regions were utilized for the estimation of transmission parameters. In comparison, the estimation of transmission rate in Models 1 and 2 exploits the data in all regions in the whole process.

#### 3.2.3 Results of Trajectory Prediction

First, we remark that in Model 5, *µ* is chosen to minimize the mean of validation errors over 100 replicas. The weighted (simply averaged) validation errors, 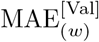 and 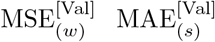 and 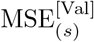, are defined as in Appendix C. We remark that the superscript ^[Val]^ refers to when the error is computed on validation data, and the subscripts _(*w*)_ and _(*s*)_ denote that the errors are the weighted and simple average of relative errors over time respectively.

The mean of weighted validation errors 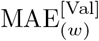 and 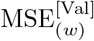 are shown in Figure 1 below, and the mean of simply averaged validation errors 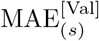 and 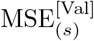 are shown in Figure A1. *µ* is chosen to be 10^2.7^, which minimizes the mean of validation errors, as can be seen from Figure 1 and Figure A1.

**Figure 1:**
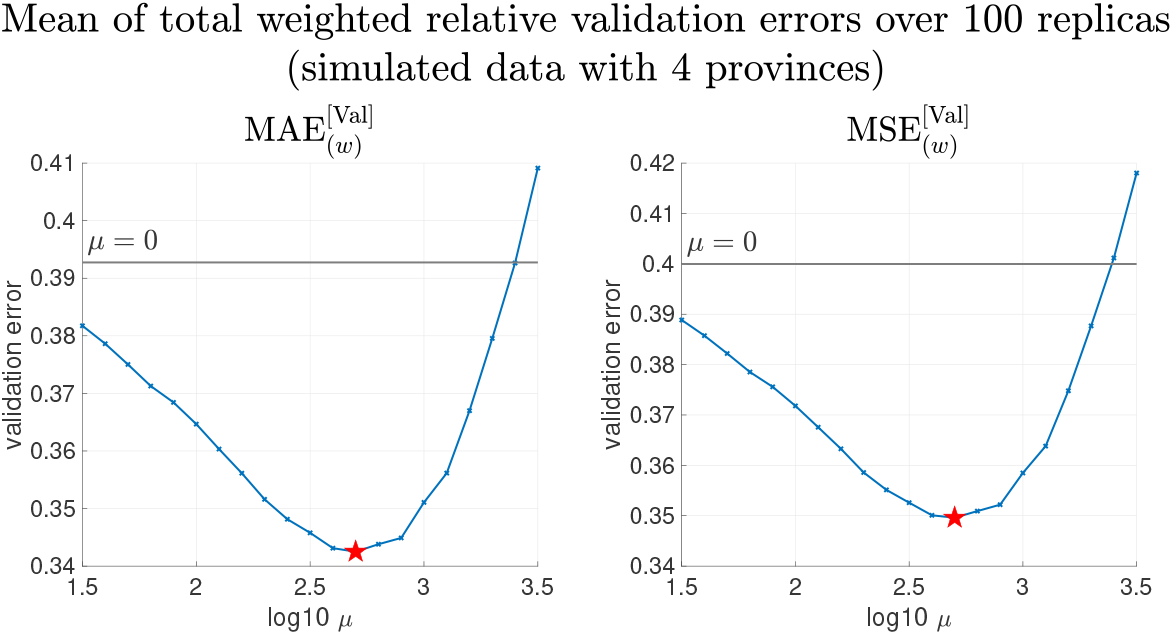
Mean of weighted validation errors 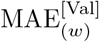 and 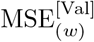 against *µ* (computed by (A5)) over 100 replicas for simulated data (four provinces). The horizontal lines show the values of the mean of the validation errors when *µ* = 0.

Then, The trajectories of a typical realization are plotted in Figure 2 and the differences between the true and fitted trajectories are shown in Figure 3. As shown in these two figures, heterogeneity helps improve the prediction of testing data more than transportation, while introducing migration without heterogeneity of parameters worsens the estimate as can also be noticed from Table 4. More explanations can be found in Section 3.2.5.

**Table 3:** Estimated *λ*_*i*_ of five models. Recall that the ground truth is *λ*_1_ = 0.5, *λ*_2_ = 0.47, *λ*_3_ = 0.4, *λ*_4_ = 0.37. Models in Column 1 are detailed in Table 2. Columns 2-4 indicate whether the model permits **migration** between provinces (for “**M**”), the **heterogeneity** of parameters (for “**H**”) and the **Graph Laplacian prior** (for “**GL prior**”), respectively. *λ*_*i*_ are inferred for each of the 100 replicas, and then the mean and standard deviation are presented in the table above. In addition, *µ* = 10^2.7^ (*σ*= 10^−6^, 10^−3^, 10^0^ respectively) in Model 5 for inference of parameters as defined in (3.1), which minimizes mean of relative validation errors over 100 replicas as shown in Figure 1 and Figure A1.

| Model | M | H | GL prior | $\lambda_1$ | $\lambda_2$ | $\lambda_3$ | $\lambda_4$ |
| --- | --- | --- | --- | --- | --- | --- | --- |
| 1 | × | × | × | $0.4458 \pm 0.0192$ | | | |
| 2 | ✓ | × | × | $0.4459 \pm 0.0192$ | | | |
| 3 | × | ✓ | × | $0.4927 \pm 0.0331$ | $0.4620 \pm 0.0370$ | $0.3988 \pm 0.0389$ | $0.3705 \pm 0.0349$ |
| 4 | ✓ | ✓ | × | $0.4980 \pm 0.0337$ | $0.4643 \pm 0.0376$ | $0.3922 \pm 0.0407$ | $0.3605 \pm 0.0366$ |
| 5 | ✓ | ✓ | ✓ ($\sigma = 10^{-6}$) | $0.4805 \pm 0.0254$ | $0.4654 \pm 0.0260$ | $0.3979 \pm 0.0256$ | $0.3879 \pm 0.0231$ |
| | ✓ | ✓ | ✓ ($\sigma = 10^{-3}$) | $0.4805 \pm 0.0254$ | $0.4654 \pm 0.0260$ | $0.3979 \pm 0.0256$ | $0.3879 \pm 0.0231$ |
| | ✓ | ✓ | ✓ ($\sigma = 10^0$) | $0.4800 \pm 0.0254$ | $0.4648 \pm 0.0260$ | $0.3972 \pm 0.0256$ | $0.3871 \pm 0.0230$ |

**Table 4:**
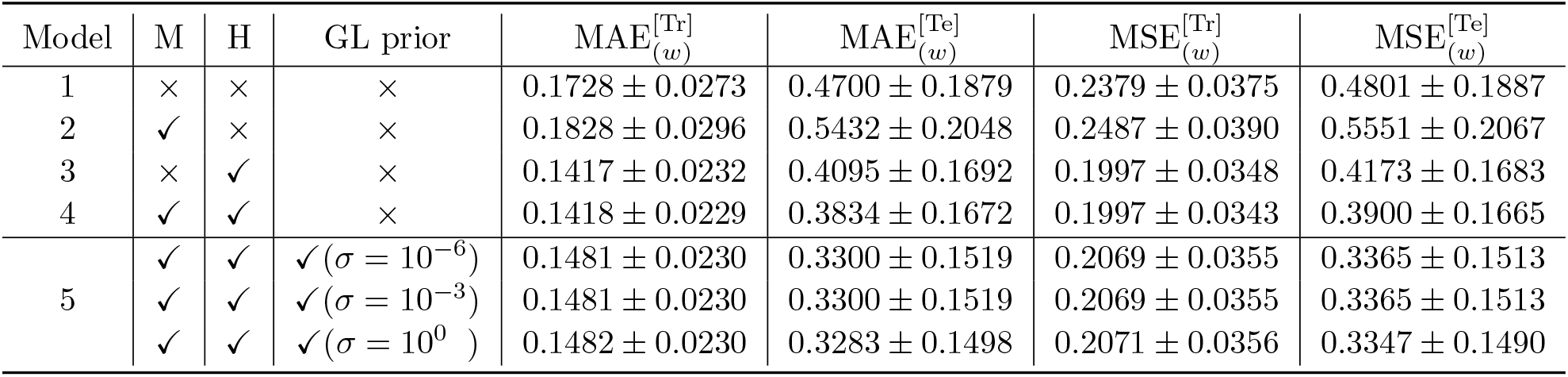
Training and testing error with standard deviation for simulated data (four provinces) of five models. The formulas of errors are detailed in Appendix C (equation (A5) or its modifications to compute testing errors). Models in Column 1 are detailed in Table 2. Columns 2-4 indicate whether the model permits **migration** between provinces (for “**M**”), the **heterogeneity** of parameters (for “**H**”) and the **Graph Laplacian prior** (for “**GL prior**”), respectively. 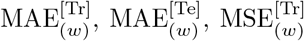 and 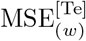 are computed for each of the 100 replicas, then the mean and standard deviation are presented in the table above. In addition, *µ* = 10^2.7^ (*σ*= 10^−6^, 10^−3^, 10^0^ respectively) in Model 5 for inference of parameters as defined in (3.1), which minimizes relative validation errors as shown in Figure 1 and Figure A1.

**Figure 2:**
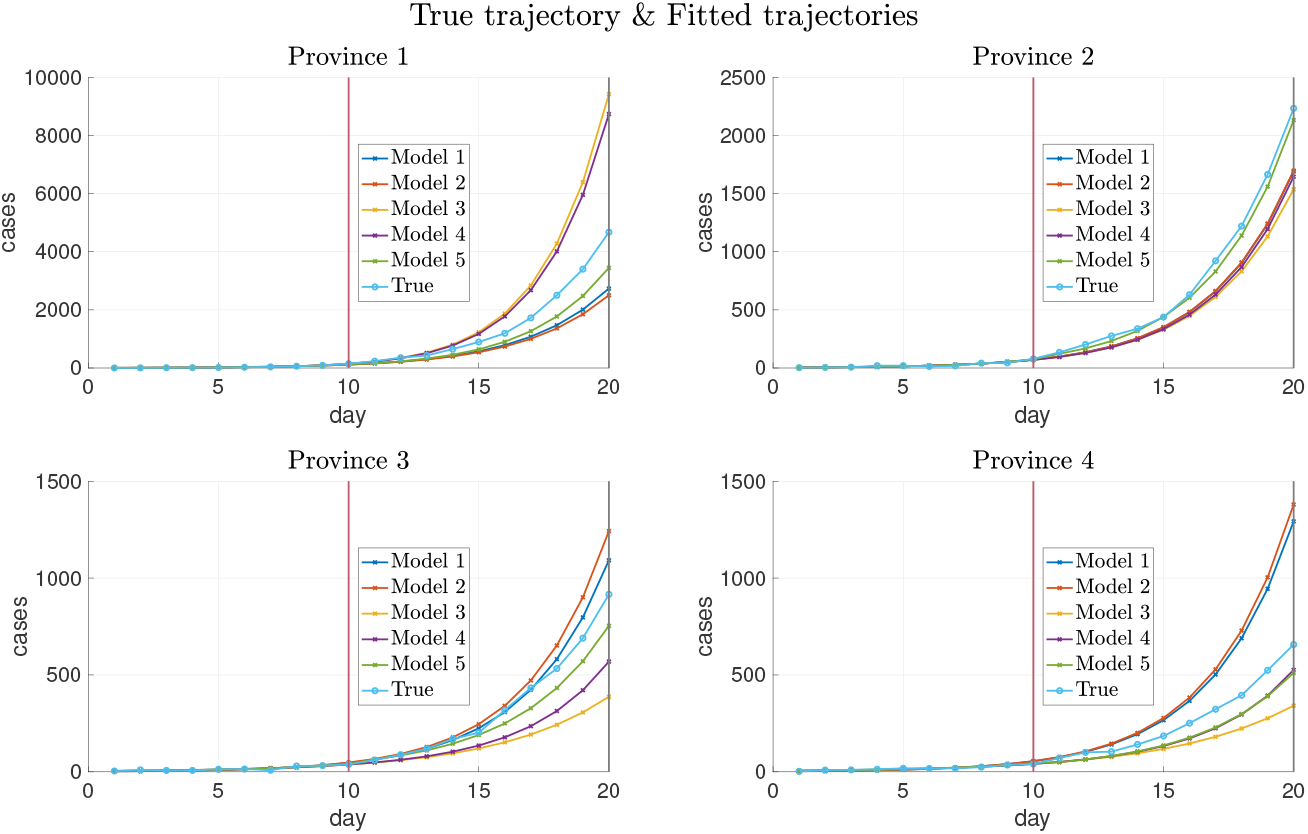
True and fitted trajectories of Models 1-5 in the four provinces. The vertical lines show the threshold *T*_*th*_ = 10.

**Figure 3:**
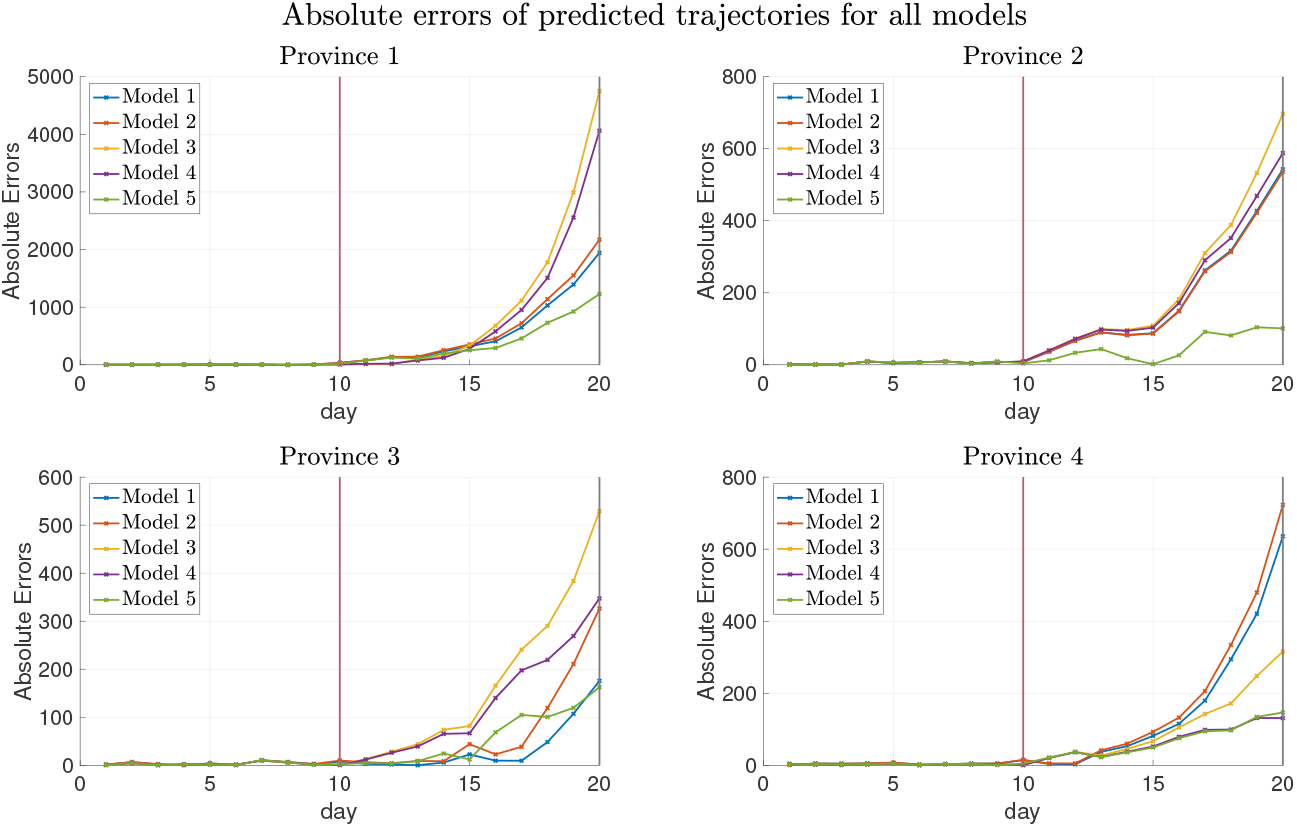
Absolute errors of fitted trajectories of Models 1-5 in the four provinces. The vertical lines show the threshold *T*_*th*_ = 10.

Additionally, Model 5 with prior distrbution based on graph Laplacian lowers the absolute errors of predicted trajectories compared with Model 4.

As shown in Figures 2 and 3, Models 1 and 2 have slightly better generalization accuracy than Model 5 for Province 3. On the one hand, for data in this replica, the estimated *λ*_3_ is 0.3860 using Model 5 and 0.4640 using Models 1 or 2 (recalling that the ground truth *λ*_3_ is 0.4). On the other hand, due to the randomness of the generated testing data, the sampled newly confirmed cases are much more than the deterministic ones in Province 3 obtained by running (2.4) with the ground truth parameters. Hence, although all these estimates of *λ*_3_ are biased from the ground truth 0.4, estimates using Models 1 and 2, which are biased up, lead to less absolute errors.

#### 3.2.4 Results of Parameter Estimation

The mean and standard deviation of {*λ*_*i*_} estimated by the five models for four provinces case are reported in Table 3. It can be observed that models allowing heterogeneity estimate parameters more accurately, and Model 5 that integrates the correlation leads to slightly better estimate for *λ*_2_. We can see that compared to Model 4, the estimates of smaller *λ*_*k*_’s (such as *λ*_2_, *λ*_3_, *λ*_4_) become larger, and the estimate of *λ*_1_ which has the largest value becomes smaller, since the graph Laplacian penalty tends to make 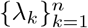 closer to each other.

Moreover, we performed a sensitivity analysis for the hyper-parameter *σ* to check that the results are robust to *σ*. The last two rows of Table 3 show the parameter estimation results for Model 5 with *σ*= 10^−3^ and 10^0^ respectively (more values of *σ* ∈ [10^−8^, 10^0^] are tested and the results are similar as well). We observe that the variation of parameters estimated by Model 5 with *σ* varying from 10^−6^ to 10^0^ does not exceed 1%. Therefore, the parameter estimation results are not sensitive to the choice of *σ* as long as *σ* is not too large.

#### 3.2.5 Further Model Evaluation

The training and testing errors, 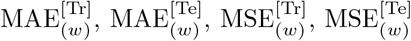, as defined in Appendix C (in the equation (A5)), are listed below in Table 4 with mean and standard deviation. We remind the readers that the superscripts ^[Tr]^ and ^[Te]^ represent the errors are computed on training and testing data respectively, and the subscript _(*w*)_ denotes that the error is weighted average of the daily relative errors over time. It can be seen from Table 4 that the presence of both heterogeneity and transportation helps reduce the training and testing errors by comparing the first four models. By comparing Model 4 and Model 5, it can be seen that using the graph Laplacian prior could further reduce the testing errors.

In addition, it can be seen from Table 4 that the errors increase greatly after transportation is included while heterogeneity remains absent. A possible explanation for this might be that without heterogeneity of parameters and transportation between provinces, the estimated *λ*_*k*_’s are lower than the true *λ*_*k*_’s for group 1, which leads to that the estimated newly confirmed cases are fewer than the true ones in group 1. For the same reason, the estimated newly confirmed cases are higher than the true ones in group 2. When the transportation is considered, more confirmed cases in group 1 are transferred to group 2 than the cases transported in the opposite direction. As a result, when the transmission parameters do not have heterogeneity, migration between provinces will worsen the prediction performance compared to the case without migration.

Furthermore, the last two rows of Table 4 report the training and testing errors for Model 5 with the same *µ* = 10^2.7^ while *σ*= 10^−3^ and *σ*= 10^0^ respectively. As a consequence of the robustness of the parameter estimation regarding *σ*, the errors of Model 5 are also robust to *σ*. The similar analysis is also performed for the other data sets and the similar results can be obtained which we do not report repetitively. Hereinafter, the results are presented with *σ*= 10^−6^.

The plots of the mean of weighted and simply averaged testing errors 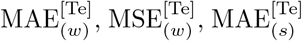 and 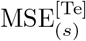 against varying *µ* are shown in Figure 4 and Figure A2 respectively. Recall that the subscripts (*w*) and (*s*) denote the weighted and simple average respectively. Note that Model 5 with *µ* = 10^2.7^, which minimizes validation errors as shown in Figure 1 and Figure A3, achieves the minimal values of testing errors 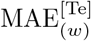 and 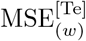 (also 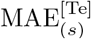 and 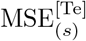). This is because validation and testing errors have the same distribution in this case. We also note that the testing error of Model 4 is nearly the same as that of Model 5 with small *µ*, since Model 4 is basically the same as Model 5 with *µ* = 0.

**Figure 4:**
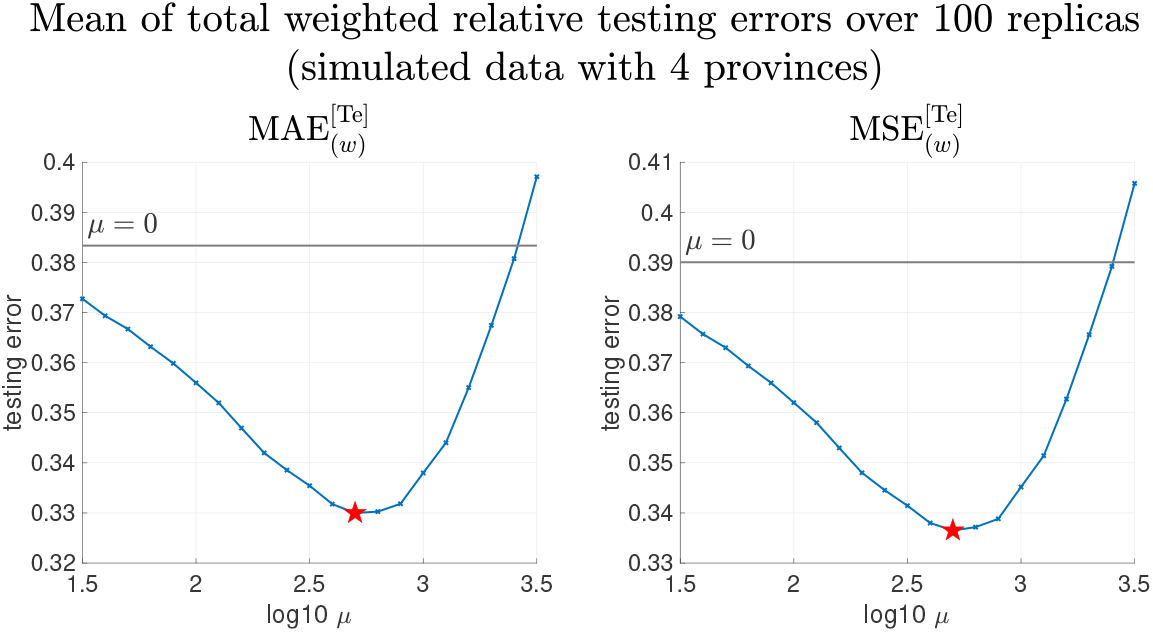
Mean of weighted testing errors 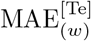 and 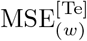 against *µ* (computed by (A5)) over 100 replicas for simulated data (four provinces). The horizontal lines show the values of the mean of the testing errors when *µ* = 0.

### 3.3 Thirty Provinces Case

In this simulation study, we present the results for the simulated data involving a larger number of regions.

#### 3.3.1 Data Description

We set the total number of regions *n* = 30, the total days considered *T* = 30, and set the threshold that separates the training and testing set as *T*_*th*_ = 10. The other prefixed parameters are listed as below:

- For *k* ∈ {1, 2, …, *n*}, *N*_*k*_(0) = 10^6^, *E*_*k*_(0) = 30, *H*_*k*_(0) = 10.
- For *l* ∈ {1, …, *T* }, *i, j* ∈ {1, 2, …, *n*} and *i* ≠*j*, (*W*_*l*_)_*ij*_ = 5 × 10^3^.
- For provinces in Group 1: *λ*_4_ = 0.307, *λ*_5_ = 0.295, *λ*_6_ = 0.303, *λ*_13_ = 0.299, *λ*_16_ = 0.296, *λ*_21_ = 0.296, *λ*_25_ = 0.294, *λ*_28_ = 0.293, *λ*_29_ = 0.305. For provinces in Group 2: *λ*_1_ = 0.416, *λ*_2_ = 0.410, *λ*_7_ = 0.405, *λ*_8_ = 0.393, *λ*_9_ = 0.400, *λ*_11_ = 0.402, *λ*_12_ = 0.406, *λ*_15_ = 0.407, *λ*_18_ = 0.395, *λ*_19_ = 0.410, *λ*_22_ = 0.396, *λ*_23_ = 0.419. For provinces in Group 3: *λ*_3_ = 0.493, *λ*_10_ = 0.512, *λ*_14_ = 0.514, *λ*_17_ = 0.503, *λ*_20_ = 0.507, *λ*_24_ = 0.498, *λ*_26_ = 0.488, *λ*_27_ = 0.495, *λ*_30_ = 0.482.

Note that the thirty provinces are randomly assigned to three groups, and we denote this ground truth partition as *P* :

- *D*_1_: Provinces 4, 5, 6, 13, 16, 21, 25, 28, 29.
- *D*_2_: Provinces 1, 2, 7, 8, 9, 11, 12, 15, 18, 19, 22, 23.
- *D*_3_: Provinces 3, 10, 14, 17, 20, 24, 26, 27, 30.

The transmission rates of the provinces in the same groups are closer as listed above. Specifically, the transmission rates of provinces in *D*_1_, *D*_2_, and *D*_3_ are randomly sampled from normal distribution with mean 0.3, 0.4, and 0.5 respectively and all with standard deviation 0.01.

#### 3.3.2 Models to Compare

We also compare the Models 1-5 listed in Table 2 as detailed in Section 3.2.2.

Note that for Model 5, the parameter *σ* in (2.8) is still fixed to be 10^−6^. When we assume that the ground truth group division *P* as listed above is known, in model performance evaluation as well as the optimization of *µ* for Model 5, by (2.11), 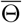 are estimated by the optimization problem in (3.2):

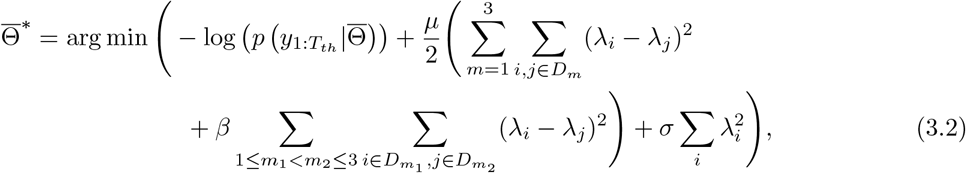

where *β* = 0.1.

Moreover, for Model 5, we also report results when there are mismatches in partition of the provinces, since in real-world applications the graph information may not be fully known. Specifically, since most of the results reported in the following sections are for Provinces 4 (in *D*_1_), 1 (in *D*_2_), and 3 (in *D*_3_), we consider the other two partitions 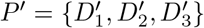 and 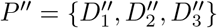, which are different from *P* in the assignments of Provinces 4, 1, and 3, and another three provinces respectively. Comparison between the results using the ground truth partition and the results of these two different kinds of mismatches may reflect the potentially different impact of these mismatches on the results. For Model 5 with partitions *P*′ or 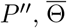 is estimated by (3.2) with *P* = {*D*_1_, *D*_2_, *D*_3_} replaced by 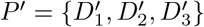 or 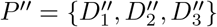.

The partition 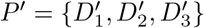 deviates from *P* in the assignment of Provinces 4 (in *D*_1_), 1 (in *D*_2_), and 3 (in *D*_3_):

- 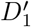: Provinces **3**, 5, 6, 13, 16, 21, 25, 28, 29.
- 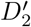 : Provinces **4**, 2, 7, 8, 9, 11, 12, 15, 18, 19, 22, 23.
- 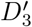 : Provinces **1**, 10, 14, 17, 20, 24, 26, 27, 30.

The partition 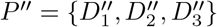 deviates from *P* in the assignment of Provinces 5 (in *D*_1_), 2 (in *D*_2_), and 10 (in *D*_3_):

- 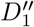: Provinces 4, **10**, 6, 13, 16, 21, 25, 28, 29.
- 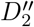: Provinces 1, **5**, 7, 8, 9, 11, 12, 15, 18, 19, 22, 23.
- 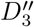: Provinces 3, **2**, 14, 17, 20, 24, 26, 27, 30.

For mismatched partitions *P*′ and *P*′′, the provinces assigned to the wrong groups are marked in bold.

We remark that the parameter inference for Models 1-4 is the same as detailed in Section 3.2.2.

#### 3.3.3 Results of Trajectory Prediction

Note that the validation and testing errors have the same distribution for the simulated data. Therefore, for Model 5 (with the ground truth partition *P*), we choose *µ* = 10^1.8^, which minimizes the mean of the validation errors as marked with red pentagrams in Figure 5 and Figure A3. The following results for Model 5 are all obtained with *µ* = 10^1.8^.

**Figure 5:**
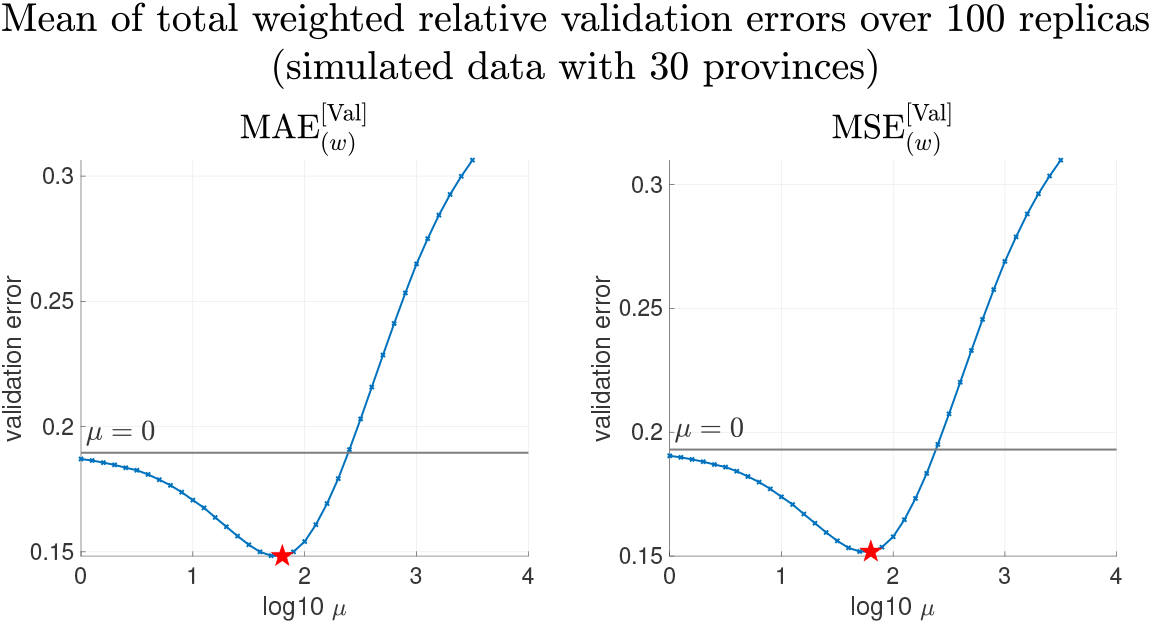
Mean of weighted validation errors 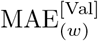 and 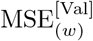 against *µ* (computed by (A5)) over 100 replicas for simulated data (thirty provinces). The horizontal lines show the values of the mean of the validation errors when *µ* = 0.

Then, we choose three provinces from the total thirty provinces (one province from each group), and plot the prediction of the trajectories and also the absolute errors from one specific replica as in Figures 6 and 7. It can be observed from Figure 7 that Model 5 (the green lines) achieve the most accurate prediction results due to the graph Laplacian regularization. Model 4 (the purple lines) also have good generalization performance, thanks to the heterogeneity and transportation involved in this model.

**Figure 6:**
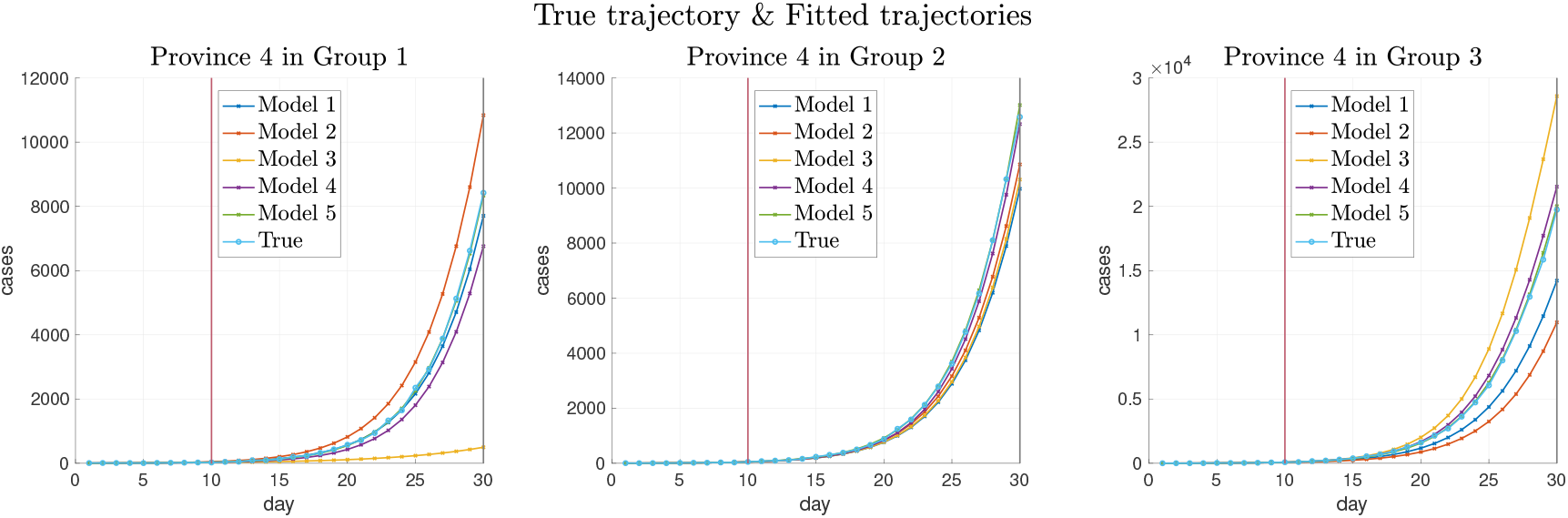
True and fitted trajectories of Models 1-5 for three provinces chosen from the total thirty provinces. The vertical lines show the threshold *T*_*th*_ = 10.

**Figure 7:**
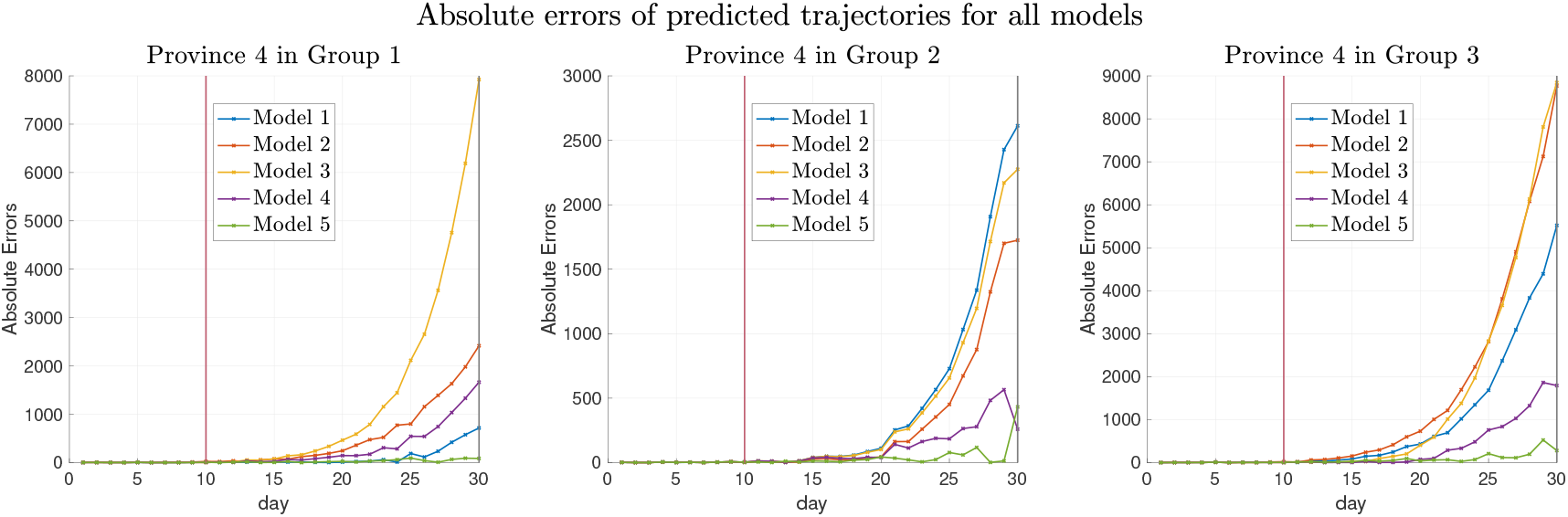
Absolute errors fitted trajectories of Models 1-5 for three provinces chosen from the total thirty ones. The vertical lines show the threshold *T*_*th*_ = 10.

We note that Model 3 also behaves much worse than the best models, especially in Provinces 4 (in *D*_1_) and 3 (in *D*_3_). In fact, the estimates for the *λ*_*k*_’s in these three provinces using Model 3 are 0.2908, 0.4198, and 0.4639, respectively, which are not much different from the ground truth values. Nevertheless, Model 3 does not involve transportation between provinces. This leads to the confirmed cases of Province 3 in *D*_3_ (which are much more than the cases of provinces in the other two groups) not output to other provinces; hence the predicted newly confirmed cases are much more than the truth. A similar situation happens in the case of Province 4 (in *D*_1_). Namely, Model 3 does not take the imported cases from the provinces in the other two groups into account, causing the predicted newly confirmed cases to be much less than the truth.

#### 3.3.4 Results of Parameter Estimation

The mean and standard deviation of *λ*_*k*_’s in Provinces 4 (in *D*_1_), 1 (in *D*_2_), and 3 (in *D*_3_) are reported in Table 5. We can still see that Models 3-5 estimate the transmission parameters more accurately than Models 1 and 2 especially for Provinces 4 and 3. In addition, the estimates by Model 5 of *λ*_*k*_ are closer than those by Model 4, due to the existence of the regularization term.

**Table 5:**
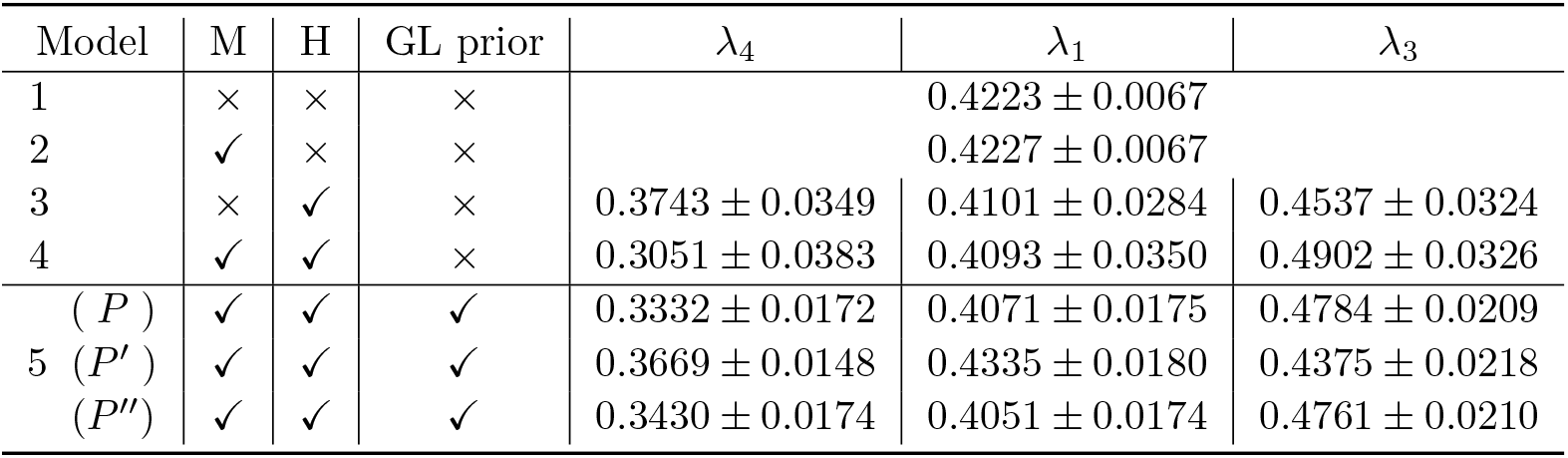
Estimated *λ*_*i*_ of five models. Recall that the ground truth is *λ*_4_ = 0.307 (in *D*_1_), *λ*_1_ = 0.416 (in *D*_2_), *λ*_3_ = 0.493 (in *D*_3_). Remarks for Columns 1-4 and estimates of *λ*_*k*_’s are the same as those in Table 3. In addition, *µ* = 10^1.8^ in Model 5 for inference of parameters as defined in (3.2), which minimizes mean of relative validation errors over 100 replicas (computed with partition *P*) as shown in Figure 5 and Figure A3. The partitions of Model 5 are *P, P*′, and *P*′′ respectively. *P* is the ground truth underlying graph structure, and *P*′ and *P*′′ are mismatched partitions introduced in Section 3.3.2.

The last three rows show the parameter estimates using Model 5 with three partitions *P, P*′, and *P*′′ respectively. Recall that *P* is the ground truth underlying graph structure, and *P*′ and *P*′′ are mismatched partitions introduced in Section 3.3.2. It can be seen that compared with the estimated parameters using Model 5 with *P*′ that differs from *P* in the grouping of Provinces 4, 1, and 3, the estimates using Model 5 with *P*′′ that differs from *P* in the grouping of Provinces 5, 2, and 10 are closer to the estimates using Model 5 with the ground truth partition *P*. Hence, the results imply that for the parameter inference of simulated data generated from a certain partition, the incorrect division causes more discrepancy in the mismatched regions when comparing the estimates using the ground truth partition.

#### 3.3.5 Further Model Evaluation

The mean and standard deviation of the weighted training and testing errors (formulas can be found in Appendix C, specifically in (A5)) are presented in Table 6. We see that Model 5 with *µ* = 10^1.8^ achieves the minimum testing errors among all the models, and hence have the best generalization performance.

**Table 6:** Training and testing error with standard deviation for simulated data (thirty provinces) of five models. The formulas of errors are detailed in Appendix C (equation (A5) or its modifications to compute testing errors). Remarks for Columns 1-4 and presented results of errors are the same as those in Table 4. In addition, *µ* = 10^1.8^ in Model 5 for inference of parameters as defined in (3.1) which minimizes mean of relative validation errors over 100 replicas (computed with partition *P*) as shown in Figure 5 and Figure A3. The partitions of Model 5 are *P, P*′, and *P*′′ respectively. *P* is the ground truth underlying graph structure, and *P*′ and *P*′′ are mismatched partitions introduced in Section 3.3.2.

| Model | M | H | GL prior | MAE<sub>(w)</sub><sup>[Tr]</sup> | MAE<sub>(w)</sub><sup>[Te]</sup> | MSE<sub>(w)</sub><sup>[Tr]</sup> | MSE<sub>(w)</sub><sup>[Te]</sup> |
| --- | --- | --- | --- | --- | --- | --- | --- |
| 1 | × | × | × | $0.1739 \pm 0.0092$ | $0.1704 \pm 0.0543$ | $0.2445 \pm 0.0135$ | $0.1744 \pm 0.0534$ |
| 2 | ✓ | × | × | $0.2427 \pm 0.0136$ | $0.3387 \pm 0.0573$ | $0.3128 \pm 0.0171$ | $0.3420 \pm 0.0566$ |
| 3 | × | ✓ | × | $0.1541 \pm 0.0082$ | $0.4920 \pm 0.0594$ | $0.2212 \pm 0.0137$ | $0.4973 \pm 0.0599$ |
| 4 | ✓ | ✓ | × | $0.1549 \pm 0.0083$ | $0.1960 \pm 0.0647$ | $0.2200 \pm 0.0139$ | $0.1994 \pm 0.0649$ |
| ($P$) | ✓ | ✓ | ✓ | $0.1648 \pm 0.0083$ | $0.1495 \pm 0.0579$ | $0.2312 \pm 0.0135$ | $0.1531 \pm 0.0570$ |
| 5 ($P'$) | ✓ | ✓ | ✓ | $0.1702 \pm 0.0084$ | $0.1612 \pm 0.0522$ | $0.2364 \pm 0.0135$ | $0.1650 \pm 0.0514$ |
| ($P''$) | ✓ | ✓ | ✓ | $0.1707 \pm 0.0085$ | $0.1648 \pm 0.0539$ | $0.2372 \pm 0.0134$ | $0.1685 \pm 0.0532$ |

The last three rows show the comparison of errors for Model 5 with three partitions *P, P*′, and *P*′′ respectively. We may see that the testing errors of Model 5 with the incorrect partitions are basically the same and larger than those with the ground truth partitions. Nevertheless, these errors are still smaller than the other baseline models. Therefore, the incorrect division might worsen the generalization performance of Model 5; however, when the division does not deviate from the ground truth much, it still behaves better than Models 1-4 with uniform prior.

Moreover, as shown in Table 6, both Model 2, which allows transportation but not heterogeneity, and Model 3, that introduces heterogeneity but no transportation, behave worse than the baseline Model 1. Model 2 having more significant testing errors is due to the same reason as the case of four provinces, as has been analyzed in Section 3.2.5. The reason for Model 3 not being able to predict well is explained in detail in Section 3.3.3.

However, we also note that Model 4, which involves both transportation and heterogeneity of parameters, has greater testing errors than Model 1 on average. This phenomenon may be partly explained by the fact that, on the one hand, there may be more infected cases imported from regions with higher *λ*_*k*_’s to those with lower transmission rates. On the other hand, although the transmission parameters are the same and all-around 0.42 in Model 1, *E*_0_ and *H*_0_ are still allowed to be spatially heterogeneous, which makes the trajectories not that far away from the ground truth. Meanwhile, though the *λ*_*k*_’s are allowed to be spatially heterogeneous in Model 4, it often underestimates *λ*_*k*_’s for provinces in *D*_1_ and overestimates *λ*_*k*_’s for provinces in *D*_3_, making the trajectories deviate from the ground truth.

Figures 8 and A4 plot the change of weighted and simply averaged testing errors 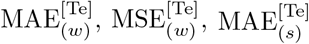 and 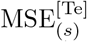 regarding *µ* (recall that the subscripts (*w*) and (*s*) denote weighted and simple average respectively). We can still observe that Model 5 with the minimizers of validation errors also has the minimal testing errors, since the errors have the same distribution for the simulated data.

**Figure 8:**
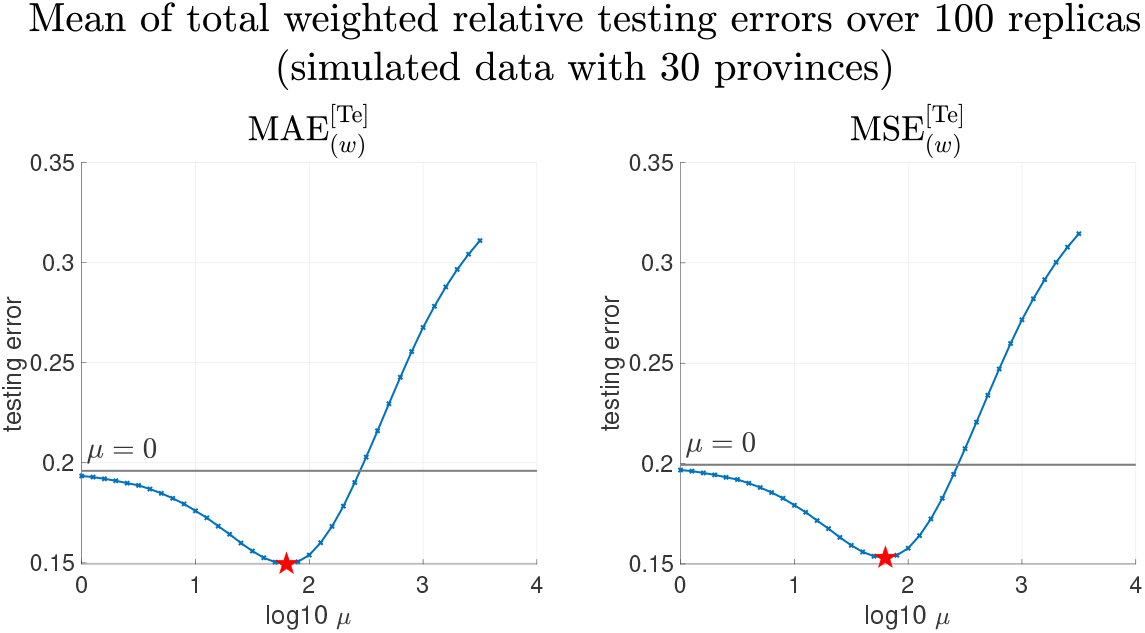
Mean of weighted testing errors 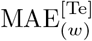 and 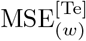 against *µ* (computed by (A5)) over 100 replicas for simulated data (thirty provinces). The horizontal lines show the values of the mean of the testing errors when *µ* = 0.

## 4 Experimental Results on COVID-19 Data

In this section, we apply our model to two real-world data sets, the COVID-19 data in China (from January to early February 2020) and Europe (from May to August 2020). We remark that the regions in Section 2 refer to provinces or municipalities for the data in China and refer to countries for the data in Europe. First, same as in Section 3, we show the predicted trajectories of the proposed model (both deterministic and stochastic ones) and the baseline models (only deterministic ones) for regions in China and Europe. We remark that regions refer to provinces or municipalities for COVID-19 data in China and countries for COVID-19 data in Europe. This shows that the proposed model can capture and further predict the trend of the true trajectories and behaves better than the baseline models, mainly due to the introduction of heterogeneity of transmission parameters and the employment of correlation between provinces or countries. After that, we show the estimate of the posterior distribution of transmission parameters in a selected province or country using the proposed model. Finally, we evaluate the models quantitatively by computing the training and testing errors as described in Appendix C, which further demonstrates the benefits of the proposed model compared to the baselines.

For the real-world data in Europe, since there might be no unique proper division, we presented the results from two different partitions, both of which are based on geographical locations of the countries. The results imply that in comparison to the simulation study where the data are generated in the artificially divided regions, the generalization performances for the real-world data of both the partitions are similar and both much better than the baseline models.

### 4.1 Data Sources

Real-world data studied in this section involve the data in China and Europe.

#### Real-world Data in China

Data in China include the number of confirmed cases and removed cases (consisting of recovered cases and fatalities) in China’s major provinces and municipalities from January 21st to March 28th (2020). These publicly available data are downloaded from the websites of the National Health Commission of the People’s Republic of China [28] and Chinese Center for Disease Control and Prevention [29]. The corresponding total population in each province or municipality is from [41].

We also utilize two transportation data sets in China extracted from Baidu Qianxi [42], lasting from January 10th to February 10th in 2020. The first data set, called as migration indexes, reflects the number of people moving out from the provinces. The second data set reflects the percentage of people moving to the corresponding destinations from their starting points. The traveling volume is estimated by combining these two data sets.

#### Real-world Data in Europe

Data in Europe include the number of confirmed and removed cases (consisting of recovered cases and fatalities) in the following 11 countries from May 1st to August 31st in 2020: Denmark, Finland, Norway, Austria, Germany, Switzerland, Italy, Spain, Belgium, France, and Ireland. The data are downloaded from [43, 13], where the data are collected from European Center for Disease Prevention and Control [30]. The data of total population in each country is obtained from [44]. During the study, we are not able to obtain suitable transportation data between these European countries needed by our model, and thus we take simplification by incorporating only spatial and temporal heterogeneity and no transportation in this experiment, details in Section 4.4.

We remark that the data in China are from the initial outbreak of the epidemic. On the contrary, the data in Europe are from when the epidemic has been ongoing about six months. The choice of the aforementioned time period of interest is to illustrate that the proposed model can predict both the case when the epidemic breaks out from some seed region and the case when the epidemic has already established sizable local spread within different regions.

### 4.2 Model Extension by Allowing Time-Varying Parameters

To capture the trend that the transmission parameters might change in real-world data due to travel restrictions or other factors, we first extend the model and parameter estimation procedure described in Section 2 so that {*λ*_*k*_} are allowed to be time-varying.

Specifically, suppose that *λ*_*k*_ is a piece-wise constant function of *t*, and 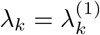 for days before some threshold *T*_*k*_, and 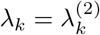 afterwards.

We denote 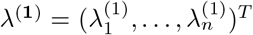 and 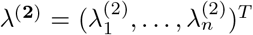 as the vectors of transmission rates in the two periods respectively. The stochastic dynamic model introduced in Section 2.2 remains unchanged except that the transmission rates will vary as time increases.

As for the parameter estimation, the first step of obtaining 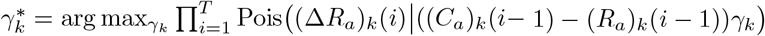 also stays the same as described in Section 2.3.

Then, when estimating 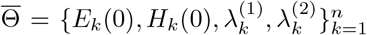 and the marginal posterior distributions of 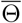, we modify the prior distribution 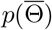 as below,

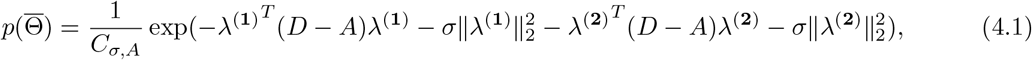

where *σ, D* and *A* have the same meaning as in (2.8), and 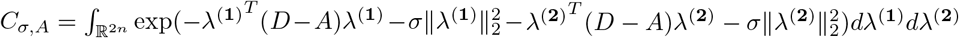 is still the normalizing constant. Consequently,

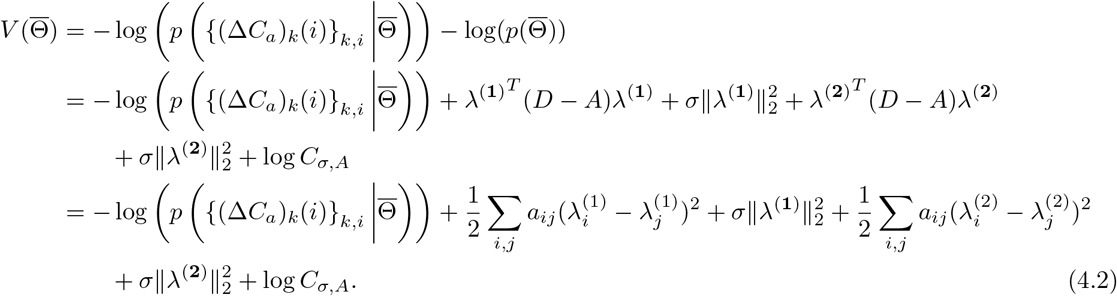

Therefore, by the definition of *a*_*ij*_ in (2.10), the estimation of 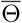 by minimizing 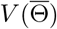 can be equivalently written as

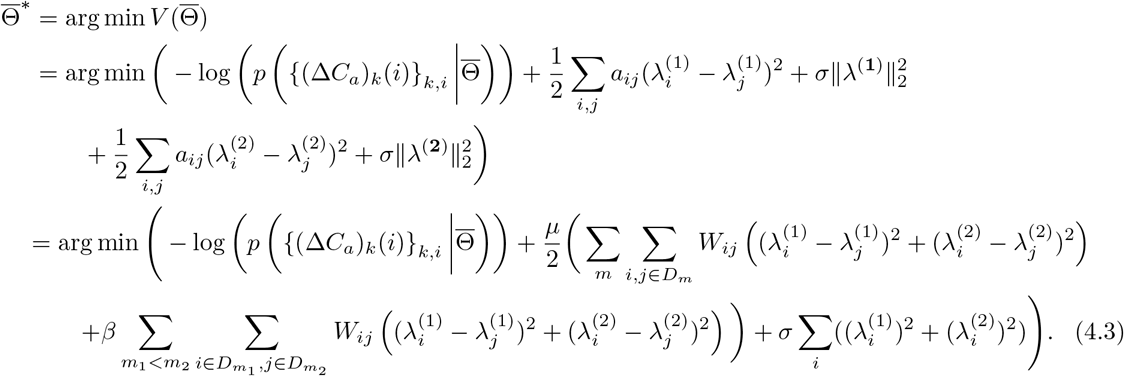

After choosing *W* and 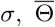 can be inferred through (4.3). Then, we could estimate the marginal posterior distributions of 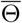 by MCMC sampling with the initial point 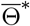.

We would like to remark that same as in the simulation study, since the proposed model depends on the penalty factor *µ* as shown in (4.3), *µ* is chosen so that the corresponding validation error achieves or is slighted higher than the minimum. Specifically, results from the multiple choices of *µ* are present, since for the real-world data, the validation data may not have the same distribution as the testing data (the choice of the training, validation, and testing sets is detailed in Appendix B and Sections 4.3.1 and 4.4.1).

### 4.3 Results for COVID-19 Data in China

#### 4.3.1 Data Description

After the selection process described in Appendix D, *n* = 21 provinces or municipalities are taken into consideration, which are Anhui, Beijing, Fujian, Gansu, Guangdong, Guangxi, Hebei, Henan, Hubei, Hunan, Jiangsu, Jiangxi, Liaoning, Ningxia, Shandong, Shan-Xi, Shanxi, Shanghai, Sichuan, Zhejiang, and Chongqing.

We set *T* = 31, from January 10th to February 10th 2020. However, we remark that only the data after January 21st are available. For measuring the fitting and generalization performance, we set the threshold *T*_*th*_ = 27 (February 5st). The data from *T*_*th*,1_ = 11 (January 21st) to the *T*_*th*_-th day (February 5st) are treated as the training set, and the data from *T*_*th*_ + 1 (February 6th) to the *t*-th day (February 10th) are treated as the testing set. Furthermore, for cross-validation of parameter *µ* of the proposed model, we choose another threshold *T*_*th*,2_ = 17 (January 27th) and another ending data *T*_2_ = 20 (January 30th). The parameters are first fitted on the data from *T*_*th*,1_ (January 21st) to *T*_*th*,2_ (January 27th), and then the validation errors are computed on the data from *T*_*th*,2_ (January 27th) to *T*_2_ (January 30th). *µ* is chosen according to validation errors.

In addition, from observation of data from January 10th to February 10th, the transmission rates in the selected provinces or municipalities change after some specific time point. Thus, we allow {*λ*_*k*_} to be time-varying for this data set with appropriate changes to the model described in Section 2, which are detailed in Section 4.2. Recall that *T*_*k*_ denotes the day when *λ*_*k*_ changes in the *k*-th region. For real-world data in China, noting that multiple containment measures, including domestic travel restrictions, contact tracing and epidemiological investigation, and even city lock down were implemented since late January/ early February, we choose *T*_*k*_ = 24 (February 2nd) for Anhui, Beijing, Henan and Hubei, and *T*_*k*_ = 20 (January 29th) for the other provinces from the initial observation of the data.

Furthermore, the data from Baidu Qianxi might not be the exact traveling volumes between municipalities and provinces. We assume that the actual traveling volume from one starting point to one destination is proportional to Baidu migration index (which reflects numbers of people departed from the stating point) and the percentage of population traveling from this origin to the destination. The corresponding scaling parameter *α* also needs to be inferred from the data for all the models.

#### 4.3.2 Models to Compare

As in Section 3.2.2, the same Models 1-5 are compared for COVID-19 data in China. Recall that Model 5 is the model proposed in this paper and the other four are the baseline models for comparison.

Note that for Model 5 with prior distribution based on graph Laplacian, the provinces are chosen to be divided into two groups, which are Hubei and other provinces except Hubei, due to the overwhelmed medical resources in Wuhan at the early stage of the pandemic [45]. For Model 5, by (4.3), the time-varying extension of (2.11) in Section 4.2, 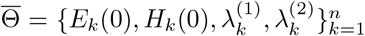 and scaling parameter *α* are estimated by the optimization problem in (4.4):

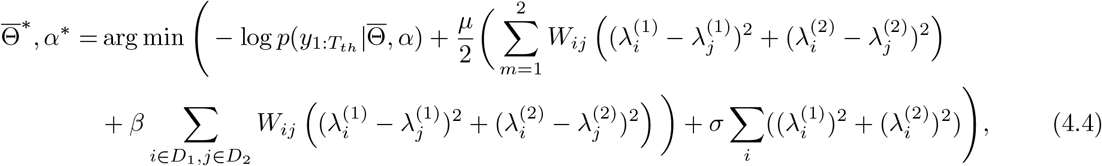

where *β* = 0.1, and 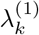 and 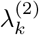 are the transmission rates in the *k*-th province before and after the *T*_*k*_-th day respectively, and *σ* is chosen to be 10^−6^. 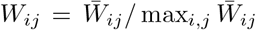, where 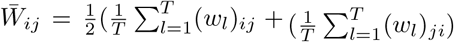 is the averaged traveling volume between the *i*-th and *j*-th province with rescaling, treated as the proximity between the two provinces.

For Models 1-4, if the model does not have heterogeneity of transmission parameters, then 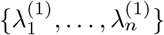 are forced to be the same and so do 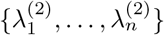, while the transmission rates in the two periods are allowed to be different; if the model does not allow transportation between provinces, then terms involving*W*_*t*_ disappear in (2.4) as described in Section 3.2.2.

#### 4.3.3 Results of Trajectory Prediction

We first remark that for Model 5, three values of *µ* are chosen, and part of the results below are present from these three choices to perform the sensitivity analysis of *µ*. The first value is *µ* = 10^2.4^, which minimizes the relative validation errors (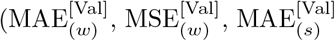, and 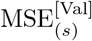), marked with black pentagrams in Figure 9 below and Figure A5. The other two values are *µ* = 10^2^ and *µ* = 10^2.7^, whose validation errors are slightly larger, marked with red and green pentagrams in Figure 9 and Figure A5 respectively. Since for the real-world data in China, the validation error does not necessarily have the same distribution as the testing error, some of the results from various choices of *µ* are presented for better comparison.

**Figure 9:**
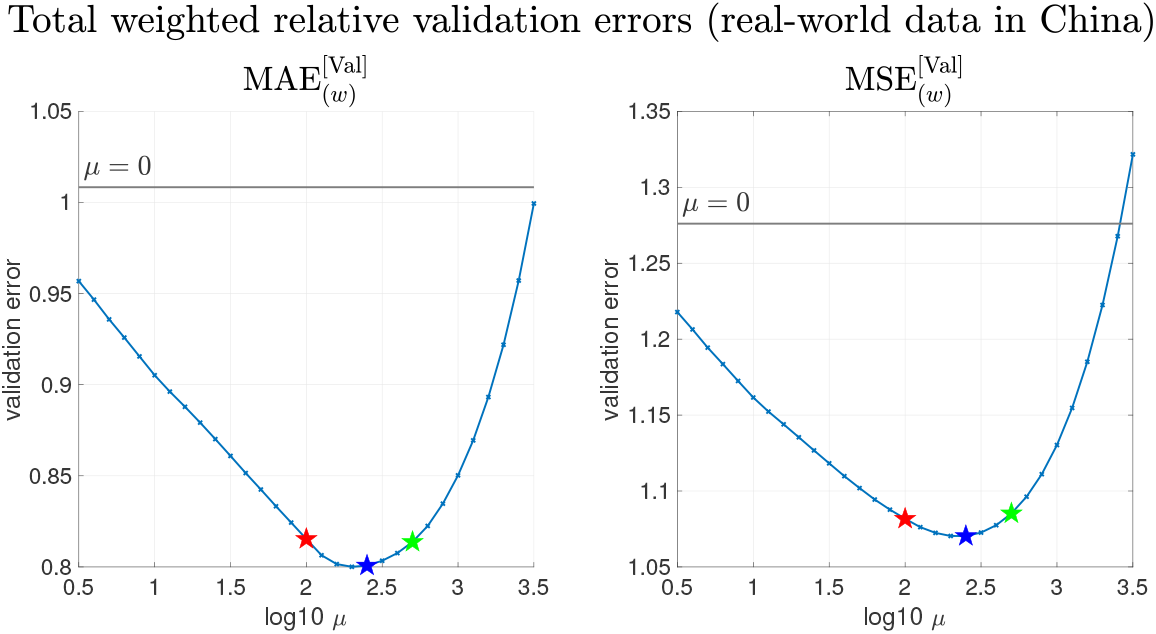
Weighted validation errors 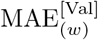 and 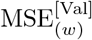 against *µ* (computed by (A5)) for real-world data in China. The horizontal lines show the values of the validation errors when *µ* = 0.

Figure 10 shows the true and predicted trajectories for newly confirmed cases in Hubei. In Figure 10,

**Figure 10:**
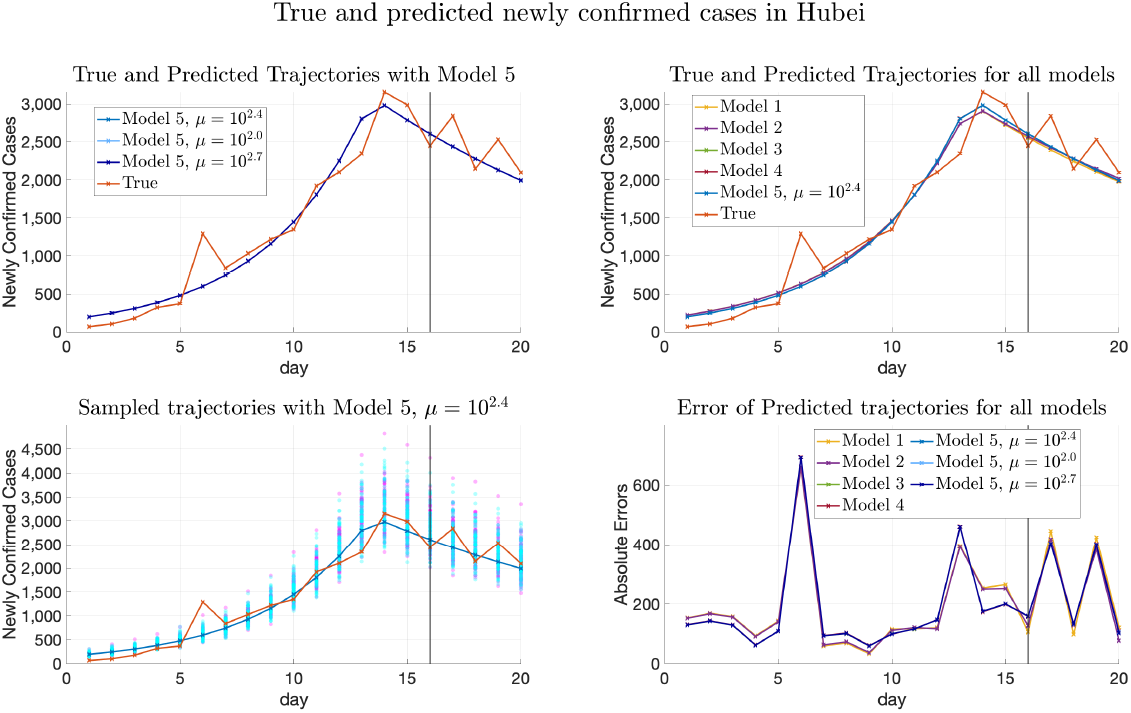
True and fitted trajectories in Hubei. The red line shows the true trajectory, the black lines show the predicted deterministic trajectories with 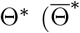 inferred using (4.4) with *µ* = 10^2.4^, 10^2^, 10^2.7^ respectively), black and red scatter plots show 100 stochastic trajectories with Θ^∗^ and sampled from the posterior distribution of 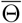 respectively (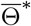 inferred using (4.4) with *µ* = 10^2.4^). The vertical lines show the threshold *T*_*th*_ = 27 for the training data and testing data.

- The red line shows the true trajectory.
- The black lines show the predicted deterministic trajectories obtained by running (2.4) with Θ^∗^ inferred using (4.4) with 10^2^, 10^2.4^, 10^2.7^ respectively.
- The black scatter plots show 100 stochastic trajectories with Θ^∗^ inferred using (4.4) with 10^2.4^
- The red scatter plots show 100 stochastic trajectories sampled from the posterior distribution of 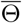, which is estimated by 5 × 10^5^ MCMC iterations staring from 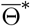 inferred using (4.4) with 10^2.4^

The deterministic trajectories obtained by the other four models and the corresponding absolute errors are also plotted for better comparison. It can be seen that all the predicted deterministic trajectories generally capture the trend of the true trajectory. Moreover, sampling trajectories from the posterior distribution of 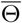 generates more randomness than sampling trajectories with Θ^∗^. Besides, it can be seen from Figure 10 that the performances of all the five models (including Model 5 with the three choices of *µ*) are similar for Hubei. This may be explained by the fact that the cases in Hubei outnumber those in other provinces or municipalities, which makes all the models tend to fit the trajectory of Hubei best.

Similarly, Figure 11 shows the predicted trajectories in Henan. As can be seen in Figure 11, unlike Hubei, heterogeneity of parameters helps the model fit and predict the trajectory of Henan in a more significant manner. When the transmission parameters in the two periods are forced to be the same in all provinces, the increment of the newly confirmed cases of the predicted trajectory is faster than the trend shown in the true trajectory in the first period. Meanwhile, the decrement of the newly confirmed cases of the fitted deterministic trajectory in the second period is also slower than the trend reflected in the true trajectory. Thus, heterogeneity helps improve the performance of fitting and predicting. Additionally, although *µ* = 10^2.4^ minimizes the validation errors, it can be seen from Figure 11 that the deterministic trajectory obtained by Model 5 with *µ* = 10^2^ achieves better performance in prediction than the one obtained by Model 5 with *µ* = 10^2.4^. Therefore, the result suggests that the choice of *µ* is not necessarily limited to the minimizer of the validation error. Instead, we may also compare trajectories with *µ* that have slightly larger validation error for possibly better generalization performance.

**Figure 11:**
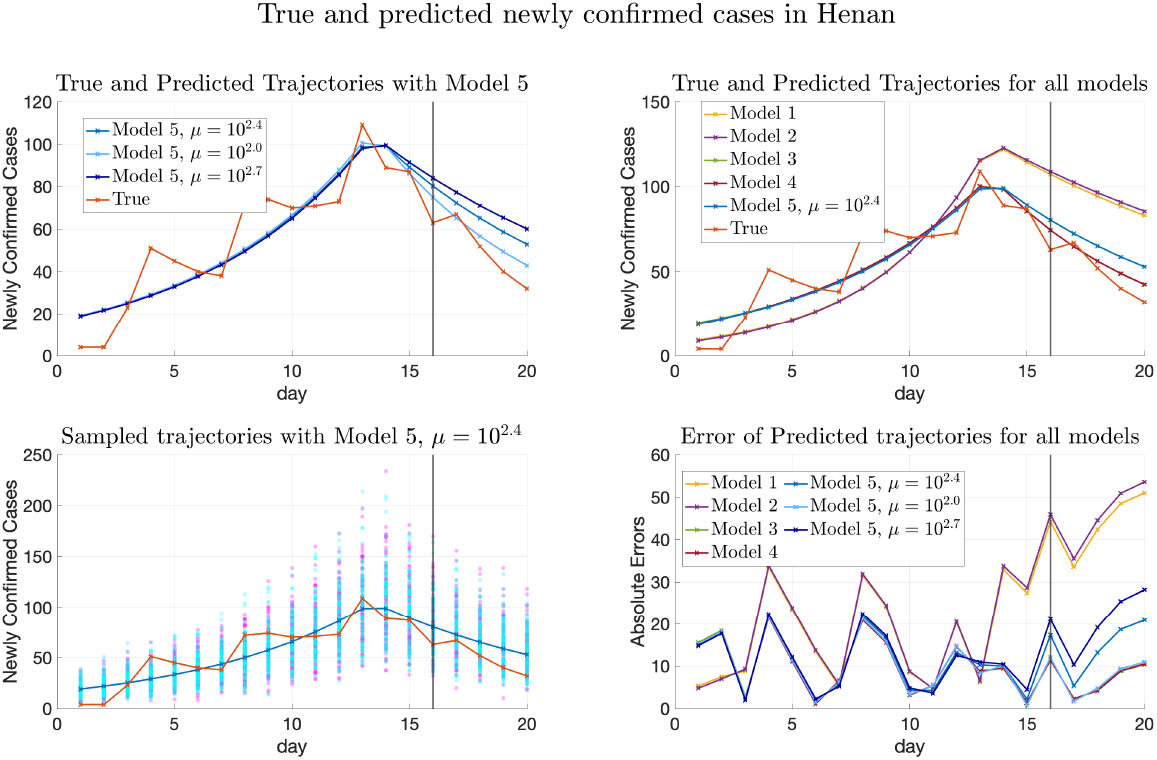
True and fitted trajectories in Henan. The red line shows the true trajectory, the black lines show the predicted deterministic trajectories with 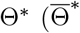 inferred using (4.4) with *µ* = 10^2.4^, 10^2^, 10^2.7^ respectively), black and red scatter plots show 100 stochastic trajectories with Θ^∗^ and sampled from the posterior distribution of 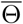 respectively (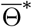 inferred using (4.4) with *µ* = 10^2.4^). The vertical lines show the threshold *T*_*th*_ = 27 for the training data and testing data.

Moreover, from Figure 12 which shows the fitted trajectories in Anhui, it can still be seen that heterogeneity helps the models predict the testing trajectory better. Furthermore, introducing correlation helps Model 5 (with *µ* = 10^2.4^) predict the trajectory marginally better than the other models.

**Figure 12:**
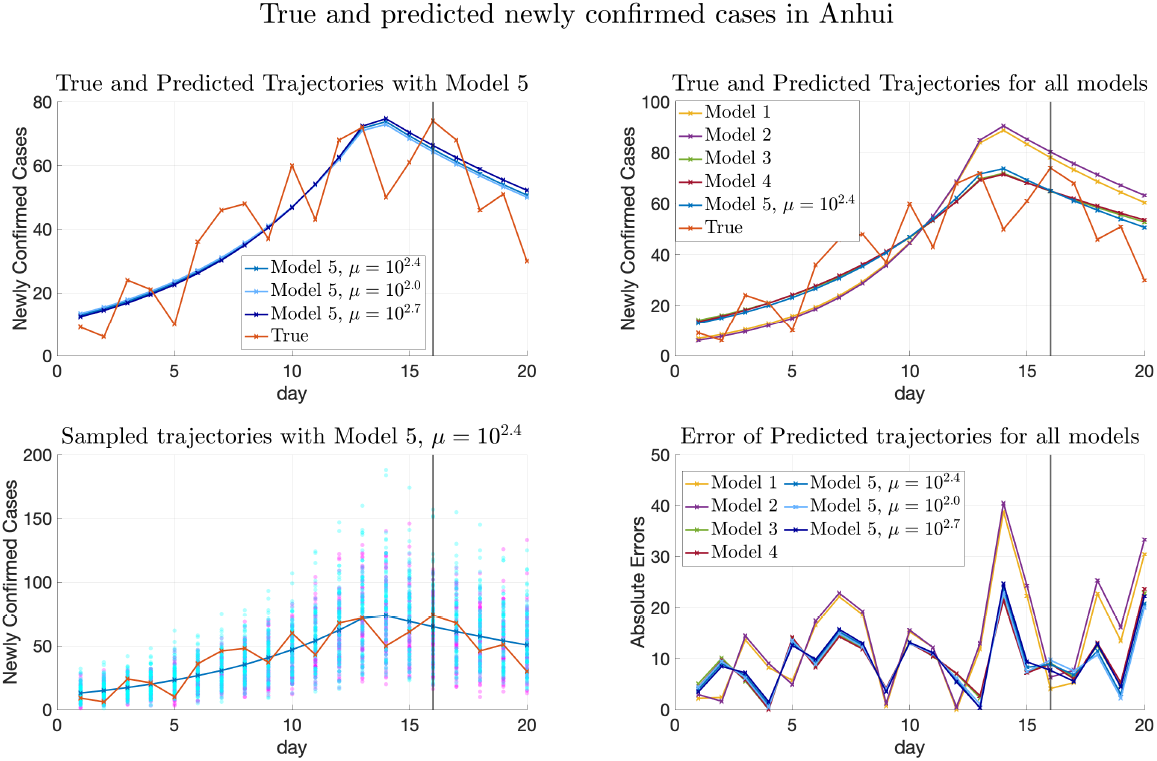
True and fitted trajectories in Anhui. The red line shows the true trajectory, the black lines show the predicted deterministic trajectories with 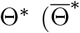 inferred using (4.4) with *µ* = 10^2.4^, 10^2^, 10^2.7^ respectively), black and red scatter plots show 100 stochastic trajectories with Θ^∗^ and sampled from the posterior distribution of 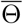 respectively (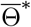 inferred using (4.4) with *µ* = 10^2.4^). The vertical lines show the threshold *T*_*th*_ = 27 for the training data and testing data.

#### 4.3.4 Results of Parameter Estimation

To illustrate the estimate of the posterior of parameters, Figure 13 shows the estimated posterior distributions of 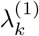 and 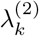 in Hubei using Model 5, in which the red lines show the inferred 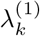 and 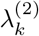 with *µ*_1_ = 10^−3.5^, and Table 7 shows the inferred 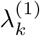 and 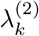 with the mean and standard deviation of their estimated posterior distributions from MCMC.

**Table 7:** Estimated paramters in Hubei. *θ*^∗^ is inferred using (4.4) with *µ* = 10^2.4^, which minimizes validation errors as shown in Figures 9 and A5. The posterior distribution is estimated by 5 × 10^5^ MCMC iterations.

| | $\theta^*$ | mean of estimated posterior | standard deviation of estimated posterior |
| --- | --- | --- | --- |
| $\lambda_{\text{Hubei}}^{(1)}$ | 0.3592 | 0.3592 | $3.932 \times 10^{-4}$ |
| $\lambda_{\text{Hubei}}^{(2)}$ | 0.0724 | 0.0722 | 0.0078 |

**Figure 13:**
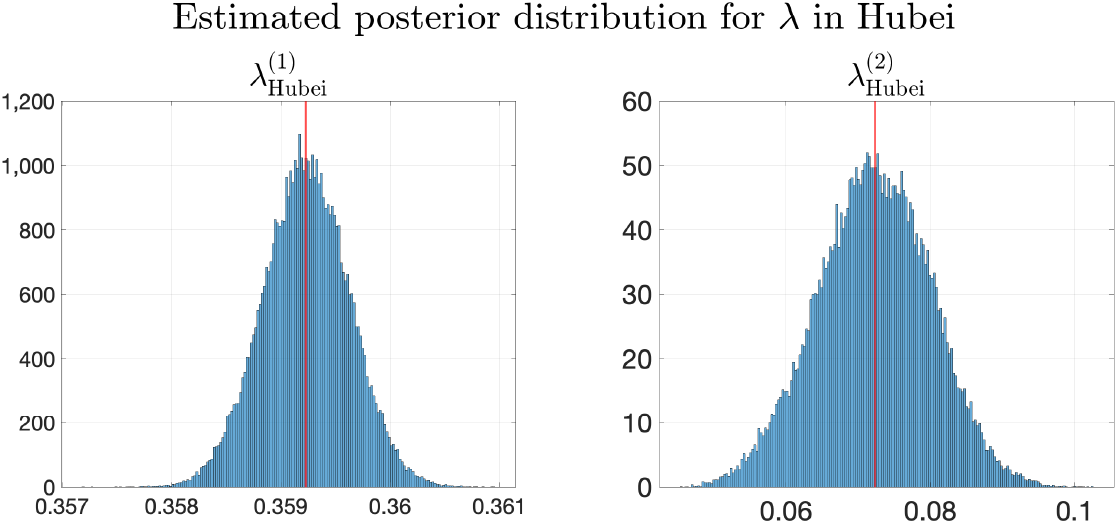
Estimated posterior distribution of *λ* in Hubei with *µ*_1_ = 10^2.4^. The vertical red lines represent the values of corresponding *θ*^∗^.

By comparing the transmission rates in Hubei in the two periods, it can be seen that the transmission rate decreases greatly after February 2nd, which indicates that the containment measures adopted in Hubei against the COVID-19 outbreak are effective.

#### 4.3.5 Further Model Evaluation

Table 8 shows the training errors and testing errors of the five models computed as detailed in Appendix C. As can be seen from Table 8, both heterogeneity of parameters and transportation between provinces help reduce the training errors and the testing errors as the simulated data case. The utilization of graph Laplacian prior helps further reduce the testing errors. Note that the testing errors of Model 4 are close to those of Model 3, which might be explained as after the traveling restrictions are imposed on January 23rd [46], transportation no longer has much influence on the spread of the epidemic.

**Table 8:**
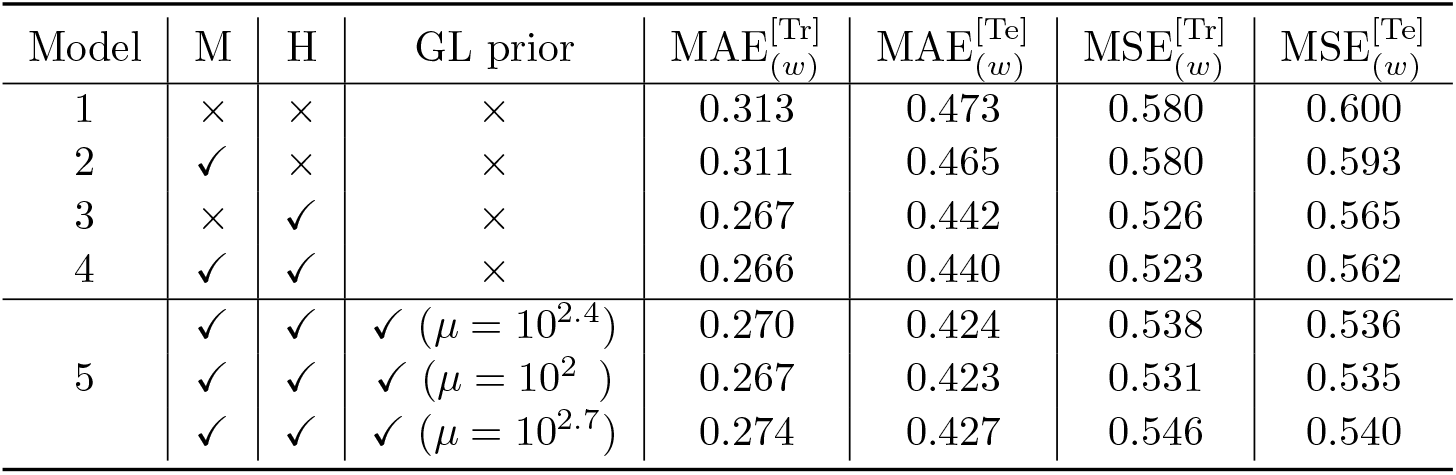
Training and testing error for real-world data in China of five models. The formulas of errors are detailed in Appendix C (equation (A5) or its modifications to compute testing errors). The remarks for Columns 1-4 are the same as in Table those in Table 4. In addition, for parameter inference with Model 5 as defined in (4.4), as shown in Figures 9 and A5, *µ* = 10^2.4^ achieves the minimal validation errors, and 10^2^ and 10^2.7^ correspond to slightly greater validation errors.

Figures 14 and A6 plot weighted and simply averaged testing errors against varying *µ* respectively. We can see from both Table 8 and Figure 14 that though *µ* = 10^2.4^ achieves the minimal validation error, it has slightly larger testing error than *µ* = 10^2^.

**Figure 14:**
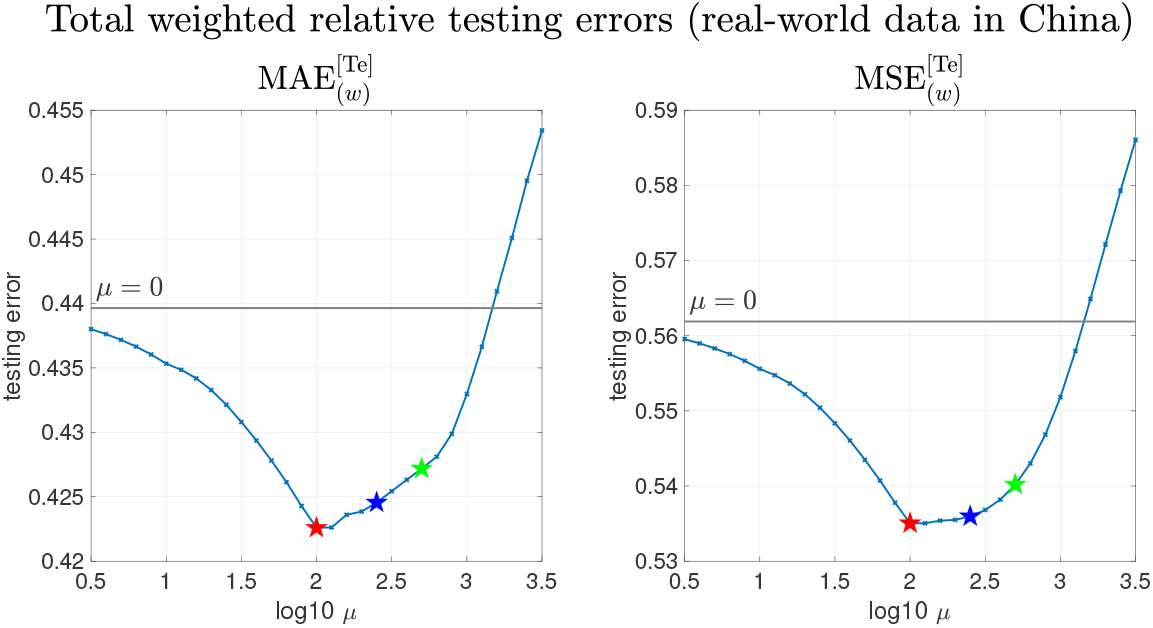
Weighted testing errors 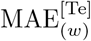 and 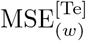 against *µ* (computed by (A5)) for real-world data in China. The horizontal lines show the values of the testing errors when *µ* = 0.

### 4.4 Results for COVID-19 Data in Europe

#### 4.4.1 Data Description

11 countries in Europe are considered in the experiments, which are Denmark, Finland, Norway, Austria, Germany, Switzerland, Italy, Spain, Belgium, France, and Ireland. Some countries such as Iceland and Luxembourg are excluded since their total population is too small, and some other countries like Sweden, Holland, and England are ignored due to that the removed cases in these countries are zero (from the data sources [43]) and thus the two-step procedure for parameter estimation in this paper may not be applied.

Since the original data oscillate much, especially over the weekends, the data are first smoothed by taking a simple moving average (which treats the unweighted average of data in 7 days around the *i*-th day as the new data point for the *i*-th day). After the smoothing, *T* = 116.

Additionally, from the observation of newly confirmed cases from May 1st to August 1st, the transmission rates change after some threshold in the selected countries in Europe. Thus, we still allow {*λ*_*k*_} to be time-varying for this data set as described in Section 4.2. *T*_*k*_ = 50 is chosen to be the day when *λ*_*k*_ changes for all countries except Norway, Italy and France. In addition, *T*_*Nowway*_ = 75, *T*_*Italy*_ = 85 and *T*_*F rance*_ = 60. Furthermore, for evaluating fitting and generalization performance, we set *T*_*th*_ = 95. For choosing *µ* (the details can be found in Appendix B.2), we set *T*_*th*,2_ = 18 and *T*_2_ = 36. Namely, the validation errors are computed by comparing the data from *T*_*th*,2_-th day to *T*_2_-th day with the predicted trajectories using parameters fitted on the data from the 1st day to *T*_*th*,2_-th day. Then, the choice of *µ* is based on validation errors.

#### 4.4.2 Models to Compare

For COVID-19 data in Europe, only three models detailed below are compared, which allow heterogeneity of parameters but do not include transportation. The last model is the proposed one in this paper, and the first two are baseline models. The transportation data needed in this model are not available to the best of our knowledge. However, the impact of transportation is expected to be less significant due to travel restrictions [46].

1’. Model with uniform prior distribution, without heterogeneity or migration.
2’. Model with uniform prior distribution, with heterogeneity but without migration.
3’. Model with prior distribution based on graph Laplacian, with heterogeneity but no migration. For Model 3’, 11 countries are partitioned into *d* = 4 groups according to geographical locations:
  - *D*_1_ (Northern Europe): Denmark, Finland, Norway;
  - *D*_2_ (Central Europe): Austria, Germany, Switzerland;
  - *D*_3_ (Southern Europe): Italy, Spain;
  - *D*_4_ (Western Europe): Belgium, France, Ireland.

We denote this partition as *P*. Since this might not be the unique appropriate partition for the *n* = 11 countries, we also present results from another partition denoted as *P*′ in the following sections. *P*′ also groups the countries that are geographically close together:

- 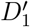: Finland, Norway;
- 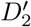: Denmark, Austria, Germany;
- 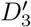: Spain, Belgium, France, Ireland;
- 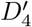: Switzerland, Italy.

The results for Model 3’ in Section 4.4.5 are presented with both the partitions *P* and *P*′.

Besides, by the same time-varying extension (4.3) detailed in Section 4.2, for Model 3’, 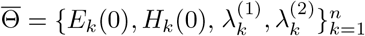 are estimated by the optimization problem in (4.5):

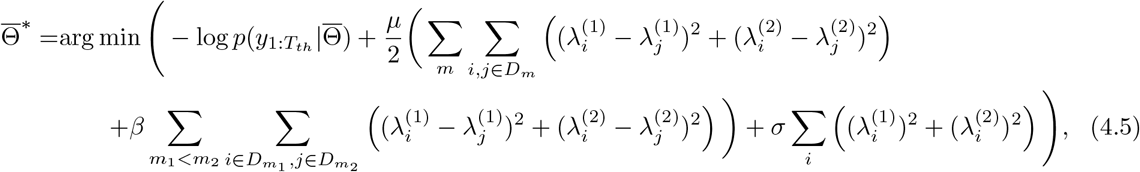

or by (4.5) with the partition *P* = {*D*_1_, *D*_2_, *D*_3_, *D*_4_} replaced by another partition 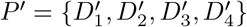. Here, *σ* is still taken to be 10^−6^, and *β* is still 0.1.

#### 4.4.3 Results of Trajectory Prediction

We still remark that for Model 3’ (with *P*), as in the case of real-world data in China, part of the results reported are from three choices of *µ* for careful consideration, since the validation error may not have the same distribution as the testing error. Note that Figures 15 and A7 plot the weighted and simply averaged relative validation errors, 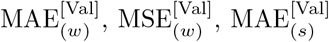 and 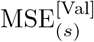 respectively. One choice is *µ* = 10^3.9^, which minimizes the validation errors (computed with *P*), marked with black pentagrams in Figures 15 and A7. The other two choices are *µ* = 10^3.2^ and *µ* = 10^5^, which correspond to slightly larger validation errors and are marked in red and green pentagrams respectively in Figures 15 and A7.

**Figure 15:**
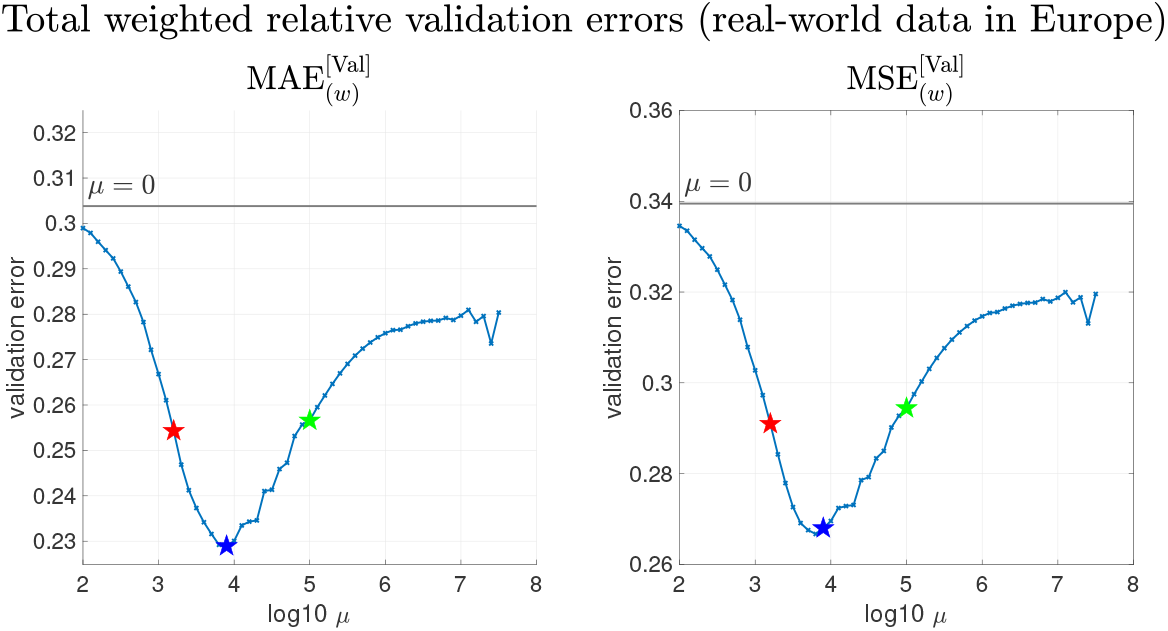
Weighted validation errors 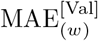 and 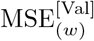 against *µ* (computed by (A5)) for real-world data in Europe. The horizontal lines show the values of the validation errors when *µ* = 0.

Figure 16, 17 and 18 present the true and predicted trajectories in Austria, Germany, and Italy, respectively. As can be seen from Figure 16, 17 and 18, heterogeneity of transmission parameters in Model 2’ (green lines) and Model 3’ (black lines) helps improve the performance of fitting and generalization. Furthermore, utilization of correlation between countries in Model 3’ further improves the prediction of the trajectories. We also note that for Model 3’, compared to the results for *µ* = 10^3.9^, which minimizes the validation errors, *µ* = 10^5^ predicts the trajectories of Austria and Italy more accurately. Hence, the results for Model 3’ are sensitive to *µ*, and it might be better to compare the trajectories from multiple choices of *µ*.

**Figure 16:**
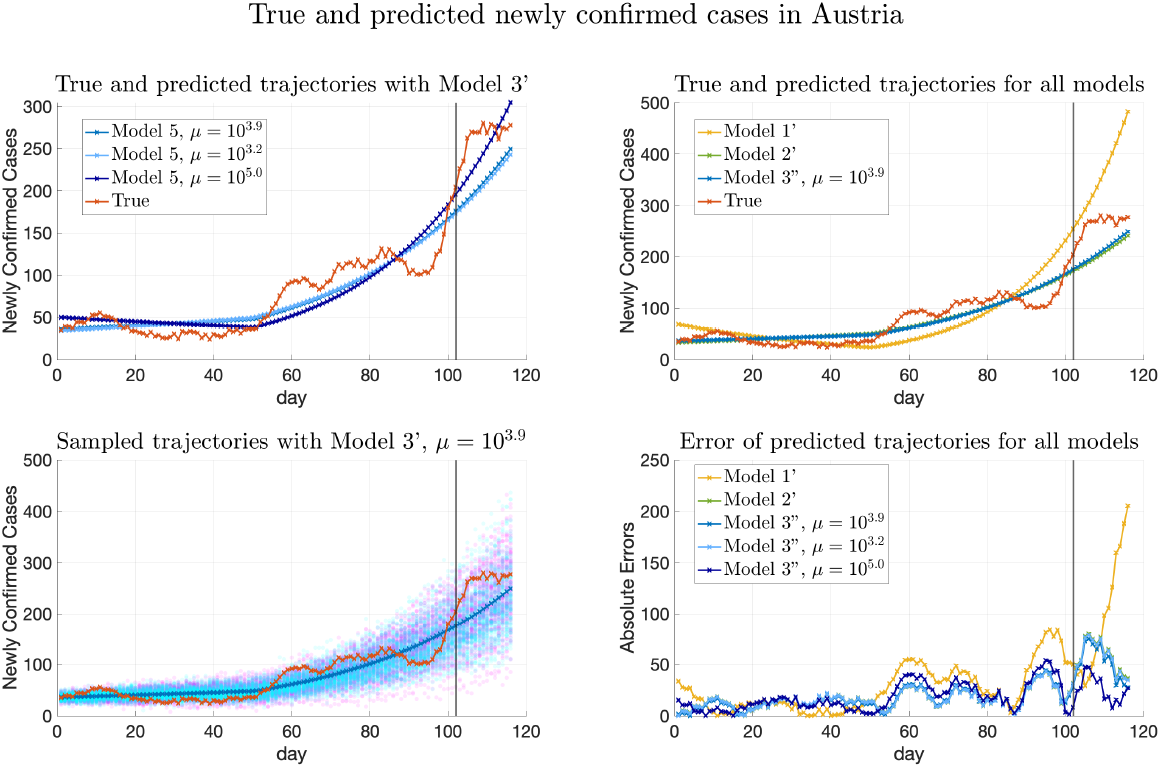
True and fitted trajectories in Austria. The red line shows the true trajectory, the black lines show the predicted deterministic trajectories with 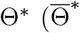 inferred using (4.4) with *µ* = 10^3.9^, 10^3.2^, 10^5^ respectively), black and red scatter plots show 100 stochastic trajectories with Θ^∗^ and sampled from the posterior distribution of 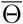 respectively (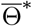 inferred using (4.4) with *µ* = 10^3.9^). The vertical lines show the threshold *T*_*th*_ = 102 for the training data and testing data.

**Figure 17:**
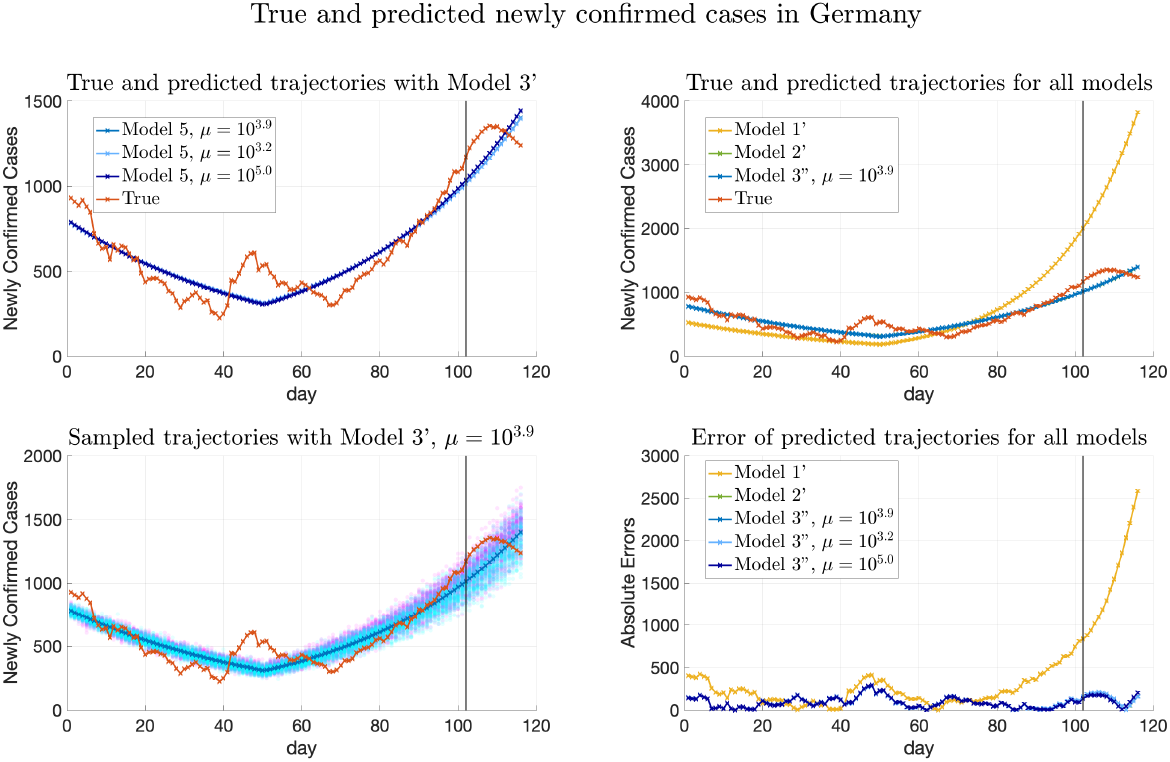
True and fitted trajectories in Germany. The red line shows the true trajectory, the black lines show the predicted deterministic trajectories with 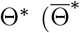 inferred using (4.4) with *µ* = 10^3.9^, 10^3.2^, 10^5^ respectively), black and red scatter plots show 100 stochastic trajectories with Θ^∗^ and sampled from the posterior distribution of 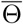 respectively (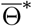 inferred using (4.4) with *µ* = 10^3.9^). The vertical lines show the threshold *T*_*th*_ = 102 for the training data and testing data.

**Figure 18:**
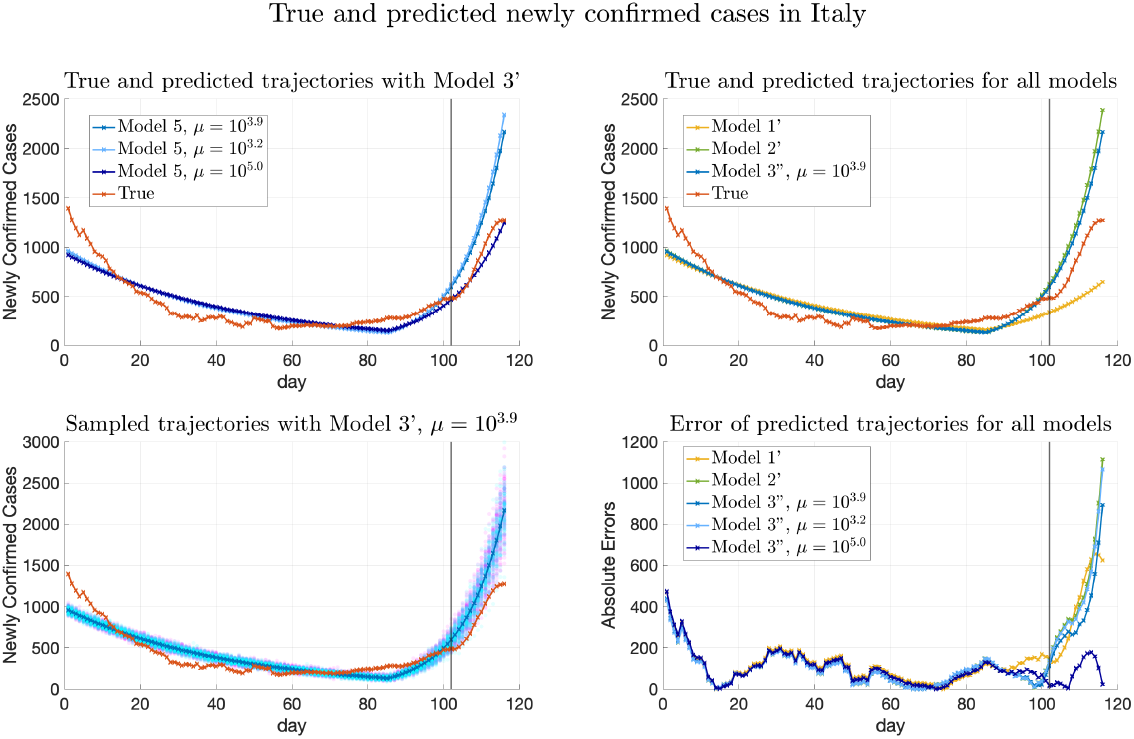
True and fitted trajectories in Italy. The red line shows the true trajectory, the black lines show the predicted deterministic trajectories with 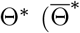 inferred using (4.4) with *µ* = 10^3.9^, 10^3.2^, 10^5^ respectively), black and red scatter plots show 100 stochastic trajectories with Θ^∗^ and sampled from the posterior distribution of 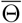 respectively (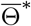 inferred using (4.4) with *µ* = 10^3.9^). The vertical lines show the threshold *T*_*th*_ = 102 for the training data and testing data.

#### 4.4.4 Results of Parameter Estimation

Same as before, Figure 19 shows the estimated posterior distributions of 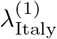 and 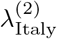 using Model 3’, in which the red lines show the inferred 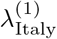 and 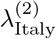 with *µ* = 10^3.9^. Table 9 shows the inferred 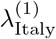 and 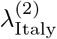 and the mean and standard deviation of their estimated posterior distribution from MCMC. The estimated 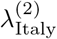 is larger than the estimated 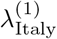, which is consistent with the trend that the newly confirmed cases in Italy first decrease and then increase.

**Table 9:** Estimated paramters in Italy. *θ*^∗^ is inferred using (4.5) with *µ* = 10^3.9^, which minimizes validation errors as shown in Figures 15 and A7. The posterior distribution is estimated by 5 × 10^5^ MCMC iterations.

| | $\theta^*$ | mean of estimated posterior | standard deviation of estimated posterior |
| --- | --- | --- | --- |
| $\lambda_{\text{Italy}}^{(1)}$ | 0.1170 | 0.1170 | $1.3794 \times 10^{-4}$ |
| $\lambda_{\text{Italy}}^{(2)}$ | 0.2336 | 0.2334 | 0.0015 |

**Figure 19:**
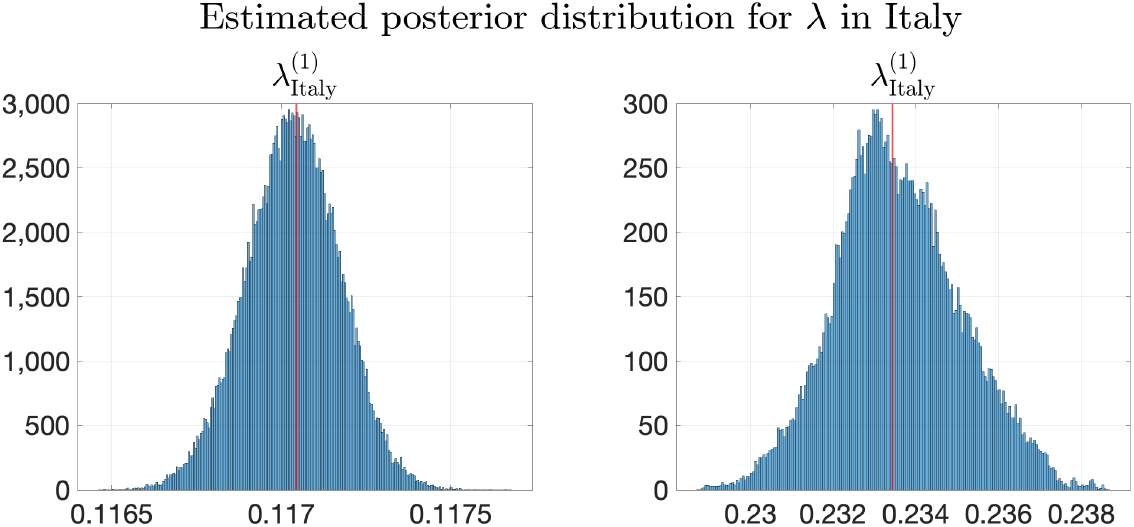
Estimated posterior distributions of *λ* in Italy with *µ* = 10^3.9^. The vertical red lines represent the values of corresponding *θ*^∗^.

#### 4.4.5 Further Model Evaluation

Table 10 shows training and testing errors of the three models. It can be seen from Table 10 that Models 2’ and 3’ have smaller both training and testing errors than Model 1’, due to the introduction of heterogeneity of transmission parameters. We can also see that compared with Model 2’, Model 3’ that adds Graph Laplacian prior has better generalization performance. Especially, Model 3’ with *µ* = 10^5^ greatly reduces the testing errors, although it does not have the minimal validation errors.

**Table 10:** Training and testing error for real-world data in Europe of three models. The formulas of errors are detailed in Appendix C (equation (A5) or its modifications to compute testing errors). The remarks for Columns 2-4 are the same as in Table those in Table 4. In addition, for parameter inference with Model 3’ as defined in (4.5), as shown in Figures 15 and A7, *µ* = 10^3.9^ achieves the minimal validation errors, and 10^3.2^ and 10^5^ correspond to slightly greater validation errors. The partitions in Model 3’ are *P* and *P*′ respectively. *P* and *P*′ are two different partitions of countries in Europe introduced in Section 4.4.2.

| Model | M | H | GL prior | $\text{MAE}_{(w)}^{[\text{Tr}]}$ | $\text{MAE}_{(w)}^{[\text{Te}]}$ | $\text{MSE}_{(w)}^{[\text{Tr}]}$ | $\text{MSE}_{(w)}^{[\text{Te}]}$ |
| --- | --- | --- | --- | --- | --- | --- | --- |
| 1' | × | × | × | 0.362 | 0.716 | 0.460 | 0.775 |
| 2' | × | ✓ | × | 0.180 | 0.531 | 0.240 | 0.601 |
| 3' ($P$) | × | ✓ | ✓ ($\mu = 10^{3.9}$) | 0.181 | 0.462 | 0.243 | 0.523 |
| | × | ✓ | ✓ ($\mu = 10^{3.2}$) | 0.180 | 0.508 | 0.240 | 0.574 |
| | × | ✓ | ✓ ($\mu = 10^5$) | 0.212 | 0.327 | 0.282 | 0.380 |
| 3' ($P'$) | × | ✓ | ✓ ($\mu = 10^{3.9}$) | 0.181 | 0.462 | 0.243 | 0.523 |
| | × | ✓ | ✓ ($\mu = 10^{3.2}$) | 0.180 | 0.510 | 0.240 | 0.577 |
| | × | ✓ | ✓ ($\mu = 10^5$) | 0.214 | 0.322 | 0.283 | 0.373 |

Recall that *P* and *P*′ are two different partitions of countries in Europe introduced in Section 4.4.2, both based on geographical locations. By comparing the corresponding results in Table 10 for Model 3’ with partitions *P* and *P*′ respectively, it can be seen that the testing errors are almost the same. This is different from the implications of the simulated data. A possible explanation for this might be that the regions in the real world have more complicated correlations than the simulated regions where clustering information is uniquely and artificially prefixed. Although reasonable groupings may not always be unique for real-world cases, the proposed model could still predict the trajectories more accurately than the baseline models.

Figure 20 below and Figure A8 plot the weighted and simply averaged testing errors (whose definitions are in Appendix C) against varying *µ* respectively, from which one may see that by imposing regularization properly through choosing a moderate *µ* helps improve the generalization performance. By comparing

**Figure 20:**
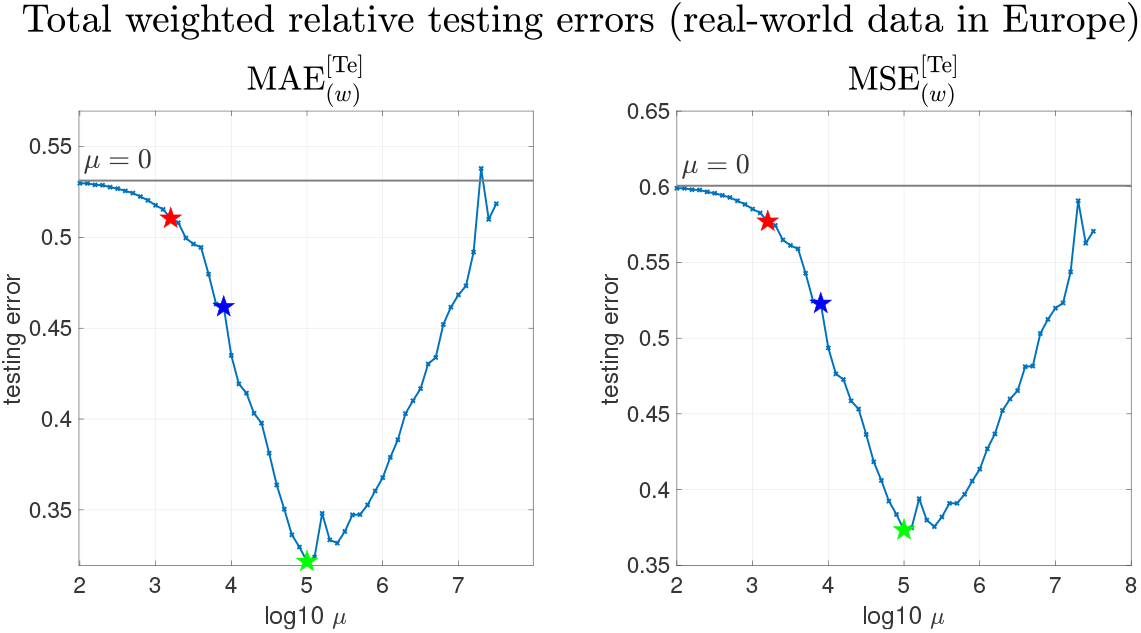
Weighted testing errors 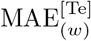 and 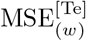 against *µ* (computed by (A5)) for real-world data in Europe. The horizontal lines show the values of the testing errors when *µ* = 0.

Figure 20 (and Figure A8) and Figure 15 (and Figure A7), we see that the change in validation and testing errors concerning increasing *µ* are not the same. This further verifies the indications from the previous findings that it would be better to examine and compare the results from multiple choices of *µ* with relatively low validation errors for Model 3’ to choose a better *µ* for the specific data sets.

## 5 Discussion

In this paper, we propose a stochastic dynamic model inspired by [1], considering the inter-district transportation and both spatially and temporally heterogeneous transmission parameters, which can model the ongoing and lasting spread of the epidemic in multiple districts. Based on this model, we also introduce a two-step procedure for estimating the parameters, which maximizes over posterior distribution with a prior distribution that utilizes graph Laplacian regularization to address the correlation between districts. Experiments on both simulated and real-world data show that compared with the baseline models, this new model with transportation, heterogeneity of parameters, and usage of the correlation in parameter inference improves the performance of fitting and generalization.

We acknowledge that there are limitations to this work. First, the stochastic dynamic model proposed in this paper might not be comprehensive enough to characterize the spread of COVID-19 in reality. For example, the model does not account for the ascertainment rate of the positive cases, the asymptomatic virus carriers, and the infectious latent period. Second, the proposed model does not consider the heterogeneity with respect to transmission risk in different populations, for example, populations with different ages, jobs, or health conditions, and needs further modification to incorporate the large-scale COVID-19 vaccination as well as waning of immunity over time. Third, the parameter inference depends on the correlation between regions determined by the clustering patterns among regions. However, such division is usually not fully known in the real-world cases, and the inference of graph structure is yet to be explored in this paper. Finally, if more complete and accurate transportation data are available, then the proposed model might predict the trend of the epidemic more accurately.

The current methods have possible extensions in the following several directions in our future study. First, the dynamic model is to be refined to be closer to reality, for example, taking the asymptomatic virus carriers and contact tracking into account and modeling the change of the transmission parameters over time in a more sophisticated way. Second, the model can be modified to accommodate the heterogeneity of various populations, including different age groups and vaccination statuses. For example, the compartments are to be further divided into subgroups, and the parameters such as transmission rates and mortality rates are allowed to be different; and the dynamic model is also to be adjusted accordingly. Third, the method could further include detecting the underlying graph structure of the regions so that the construction of the graph Laplacian matrix could be more self-contained and systematic. At last, the method could be further used to assess the influence of containing measures taken by different countries, for example, by adjusting the traveling volume to make it different from the actual transportation data and then analyzing the corresponding influence on the size and trend of the pandemic.

## Data Availability

All data produced are available online at https://github.com/Yixuan-Tan/Statistical_Inference_Using_GLEaM_with_Spatial_Heterogeneity_and_Correlation

## Appendix A Review of SEIR Model and the GLEaM Model

In this section of Appendix, we provide a more detailed introduction to the SEIR Model and the GLEaM Model so as to provide a background of our study and augment the main text.

To depict the evolution of the epidemics, [19] proposed the celebrated Susceptible-Infected-Removed (SIR) model and characterized the development of the pandemic with a deterministic ordinary differential equation (ODE). There are many extensions of the SIR model, including the Susceptible-Exposed-Infected-Removed (SEIR) model for diseases with a latent period, the Susceptible-Infected-Susceptible(SIS) model for diseases that do not gain immunity after recovery, etc.

These deterministic transmission models are constructed certain assumptions, including that the population is large, closed, and homogeneous. Due to the random nature of the transmission process, many stochastic dynamic models are developed [47, 48, 49]. Under certain rather generalized conditions, the deterministic models can be seen as the mean-field equations of the corresponding stochastic processes. However, this approximation may not hold when the size of the outbreak has not grown up to the same order of the total population, which is the case in many applications [50]. More details can be found in[34] and the references therein.

The Global Epidemic and Mobility (GLEaM) model proposed in [1] used a meta-population scheme which balanced between the agent-based stochastic models and the deterministic compartmental models. Specifically, [1] adapted a high-resolution population database that divided the surface of the earth with cells of 15min × 15min of arc, and then used Voronoi tessellation to assign each cell to one of the major airports around the world. The obtained subdivisions were then called subpopulations.

The stochastic dynamic in the subpopulations was then coupled with two layers of mobility flows apart from the infection dynamic within each subpopulation. The first layer was the worldwide airport network between the airports in the subpopulations, which could be seen as a weighted graph whose edges represented the number of passengers between each pair of airports. This layer was integrated into the model through stochastic transportation between subpopulations. The second layer was the commuting network that connected subpopulations graphically close. This layer was integrated through being used to compute the effective population and infection in each subpopulation. More details can be found in [1].

## Appendix B Data Splitting and Choice of Hyper-parameters

In this section, we first specify the choice of training, validation, and testing data, and explain the choice of the hyper-parameter *µ* through cross-validation in more detail for simulated and real-world data in Section B.1. Then, we detail the choice of the algorithmic hyper-parameters in Section B.2, including *σ* in (2.8), the penalty factor *µ*, the parameter *β* ∈ (0, 1) in (2.10), and partition of regions.

### B.1 Construction of Training/Testing/Validation Data

In this section, we detail how the training, testing, and validation data are chosen for the simulated and real-world study in Sections B.1.1 and B.1.2 respectively. Then, we also explain how the fitted/predicted trajectories are computed for the training/testing/validation data. The corresponding errors are calculated as the (weighted) average of daily relative errors for the fitted/predicted trajectories compared with the ground truth data.

We remark that the choice of training and testing data are for evaluating the model performance through training and testing errors. In addition, choosing validation data aims to help select hyper-parameters in the proposed model through validation errors, as will be mentioned in Section B.2. The details of computation of errors can be found in Section C.

#### B.1.1 Simulated Data

For simulated data, three random trajectories are sampled independently according to the stochastic model with prefixed parameters in one replica. For some threshold *T*_*th*_, data from the first *T*_*th*_ days of one trajectory are treated as the training data 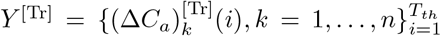, data from the last *T* − *T*_*th*_ days of the second trajectory are treated as the validation data 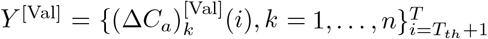, and data from the last *T* − *T*_*th*_ days of the last trajectory are treated as the testing data 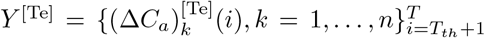. This guarantees that the training and validation data are independent from the testing data and that validation data have the same distribution as testing data.

The fitting performance is assessed by comparing the training data *Y* ^[Tr]^ with the fitted deterministic trajectory 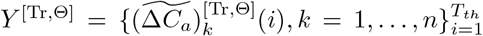 is obtained by integrating ODE system (2.4) for *T*_*th*_ days with parameters 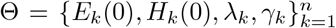 inferred by the corresponding model. The model can be either any baseline model or the newly proposed model.

Similarly, the generalization performance is assessed by comparing the testing data *Y* ^[Te]^ with the predicted deterministic trajectory 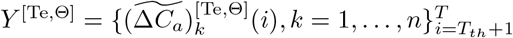. *Y*^[Te,Θ]^ is computed by running (2.4) for *T* days with parameters 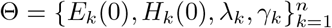 and then taking out the data of the last *T* − *T*_*th*_ days.

Moreover, to help determine the hyper-parameter in the proposed model, we evaluate the model’s performance with parameters fitted using training data on the validation data set. Specifically, for a given hyper-parameter, we first fit the parameters 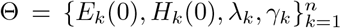 by the training data *Y* ^[Tr]^ and (2.11). Then, we compute the predicted deterministic trajectory for the validation data, 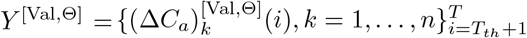, by integrating ODE system (2.4) for *T* days with the fitted Θ_*µ*_ and taking out the data of the last *T* − *T*_*th*_ days.

#### B.1.2 Real-world Data

For real-world data, only one observable trajectory is available, thus the training and testing data (*Y* ^[Tr]^ and *Y* ^[Te]^) are the two parts of the true trajectory split by the *T*_*th*_-th day. Moreover, the validation data *Y* ^[Val]^ are part of the training data *Y* ^[Tr]^, lasting from the *T*_*th*,2_-th day to the *T*_2_-th day, for some other threshold *T*_*th*,2_ and end point *T*_2_.

In contrast to the case for simulated data, for real-world data, the training data and validation data are not independent from the testing data. Instead, to measure the fitting and generalization performance for some model with the fitted parameters Θ, a deterministic trajectory for all the *T* days is computed with Θ, and then the data for the first *T*_*th*_ days and for the last *T* − *T*_*th*_ days are treated as *Y* ^[Tr,Θ]^ and *Y* ^[Te,Θ]^. Moreover, to choose hyper-parameters for the proposed model in this paper, we first fit the parameters Θ with (4.3) using the the first *T*_*th*,2_ days of training data and a given hyper-parameter. Then, we compute the predicted validation trajectory for this hyper-parameter, *Y* ^[Val,Θ]^, as the last *T*_2_ − *T*_*th*,2_ days of the deterministic trajectory of total *T*_2_ days determined by (2.4) with the parameters Θ.

We remark that for real-world data, *T*_*th*_ should not be earlier than the maximum of *T*_*k*_, which is the day that *λ*_*k*_ changes in the *k*-th region, ∀*k* = 1, …, *n*. Moreover, we choose *T*_*th*,2_ such that it is earlier than the minimum of *T*_*k*_ due to that the time span of the real-world data is not long enough especially the data in China.

### B.2 Choice of Hyper-parameters

It can be seen from Section 2.3 and Algorithm 1 that there are several hyper-parameters that determine the model proposed, including *σ* in the prior distribution (2.8) along with the factor *µ* for graph Laplacian penalty, *β*, and partition of regions that all decide the affinity *A* through (2.10). We remark on the choice of these hyper-parameters as follows.

- *σ*: the factor for regularization on *l*_2_ norm of the vector(s) of transmission parameters, appearing in both (2.11) and (4.3). In this paper, *σ* is chosen to be 10^−6^ for all the data sets. We present the results of sensitivity analysis performed for *σ* on the simulated data with four provinces in Section 3.2. The results are not sensitive to the choice of *σ* so long as *σ* is not too large. Similar patterns show for other data sets, thus details are not reported repetitively and the choice of *σ* is fixed as 10^−6^ throughout the experiments.
- Partition of regions: the partition determining construction of the affinity matrix *A*, such that correlation between regions from the same group in the partition is more addressed. For simulated data, experimental results are mainly reported with underlying graph information known. For real-world data, the partition of provinces in China is based on the medical overrun in Hubei at the initial outbreak of the epidemics, and the partition of countries in Europe is grounded on the geographical locations. Moreover, to test whether the model works when the graph information is not fully known, we also present results from mismatched/alternative partitions for simulated/real-world data in Section 3.3 and 4.4. The results show that the proposed model still predicts more accurately than the baseline models if the mismatch is not too serious (for the simulated data) or the reasonable partition may not be unique (for the real-world data).
- *µ*: the penalty factor of the graph Laplacian regularization, appearing in both (2.11) and (4.3). *µ* is chosen through cross-validation. Specifically, we separate out some validation data, then fit parameters Θ_*µ*_ for each *µ* and compute validation errors by comparing the actual validation trajectories with the predicted ones with fitted Θ_*µ*_. Then, the choice of *µ* depends on the validation errors. In the Sections B.1.1 and B.1.2 above, we have specified how to choose validation set for simulated and real-world data in detail respectively. In addition, the formulas of validation errors can be found in Section C. For the case of simulated data, validation errors are identically distributed as testing errors, which are used to evaluate the prediction performance of models, *µ* is chosen to minimize the validation errors. On the contrary, for real-world data when validation errors may have different distribution from testing errors, results from multiple choices of *µ* with relative small validation errors are presented. Details of results are in corresponding sections of experimental results.
- *β*: the parameter taking values in (0, 1) that reduces the correlation between inter-group regions through (2.10). In this paper, *β* is fixed to be 0.1 for all the experiments. We remark that other reasonable choices of *β* ∈ (0, 1) would also lead to similar results, which might not be exactly the same as those with *β* = 0.1 though. *β* can also be chosen through cross-validation, similarly to the choice of *µ*, and we omit the details in this paper.

## Appendix C Model Evaluation Metric

In Section B, we have specified the choice of ground truth training/testing/validation data and explained how they are estimated using models for both simulated and real-world data.

In the current subsection, we define the training, validation, and testing errors for model evaluation and help choose hyper-parameters of the model. Specifically, we introduce the definitions of mean relative error (which will be abbreviated as MAE hereinafter) and root mean squared relative error (which will be abbreviated as MSE for short), simply averaged or weighted.

In the paper, the superscripts ^[Tr], [Val]^, and ^[Te]^ refer to when the error (MAE and MSE) is computed on the training, validation and testing data respectively. We thus introduce the definition of error here omitting the superscript for simplicity.

We compute the error over the days from *i* = 1 to *T*_*e*_. Given the data of true daily confirmed cases Δ*C*_*a*_ and those estimated from the model 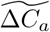 we define

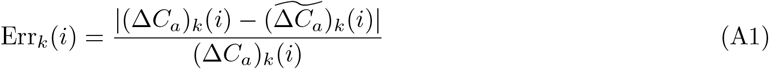

as the relative error in the *k*-th region on the *i*-th day. Then, the total error for the *k*-th region are the weighted sum of 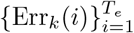. We use weighted average in computing the error, and adopt the following two types of weights *α*_*k*_(*i*) cross region *k* and time *i*. The first type of weight, denoted with subscript _(*w*)_, is defined as

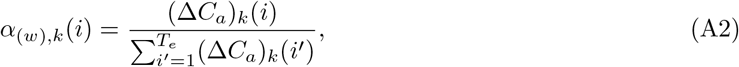

which share the same trend of increasing or decreasing with the newly confirmed cases in the *k*-th region and give larger weights to errors with more cases. The second type of weight conducts simple average over time, namely,

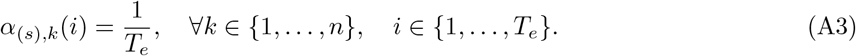

With the two types of weights (A2) and (A3), we define the weighted MAE and MSE for the *k*-th region as

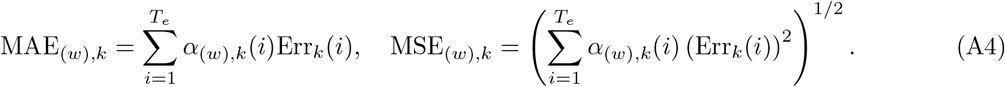

The errors weighted by *α*_(*s*),*k*_(*i*) are defined similarly and denoted with subscript _(*s*)_ instead. Note that, strictly speaking, the error for each day and region before taking average is a relative error, c.f. (A1). We use the name ‘MAE’ and ‘MSE’ following the convention.

The definition (A4) is for each region *k*. The total error over all regions is defined by taking an average over regions, that is,

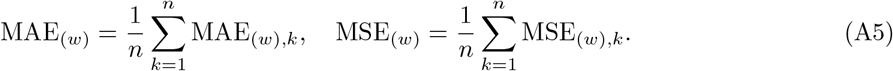

The simple-average weight counterparts MAE_(*s*)_ and MSE_(*s*)_ are defined in the same way from MAE_(*s*),*k*_ and MSE_(*s*),*k*_ respectively.

## Appendix D Preprocessing of COVID-19 data in China

Before the data is analyzed, an initial screening over provinces is applied to the COVID-19 data in China. In our study, we find this preprocessing step necessary for stability of inferring the model parameters.

Specifically, only the provinces or municipalities in China that satisfy the following conditions are considered:

1. The accumulated confirmed cases by February 10th 2020 are above 50. If the accumulated confirmed cases is too small, the newly confirmed cases touch zero frequently and the compartment model considered in this paper may not be appropriate to characterize the spread of COVID-19 in such region.
2. The estimated removal rate *δ*_*k*_ is not less than 10^−10^. If the newly removed cases are too small, then the two-step procedure proposed in this model may not estimate the parameters appropriately.
3. The change of transmission parameter occurs no later than February 2nd 2020. Note that the data in China last to February 10th 2020, and the training data end on February 5th 2020. If the changing point is too late, the training data could not capture the transmission parameter in the second period.

**Figure A1:**
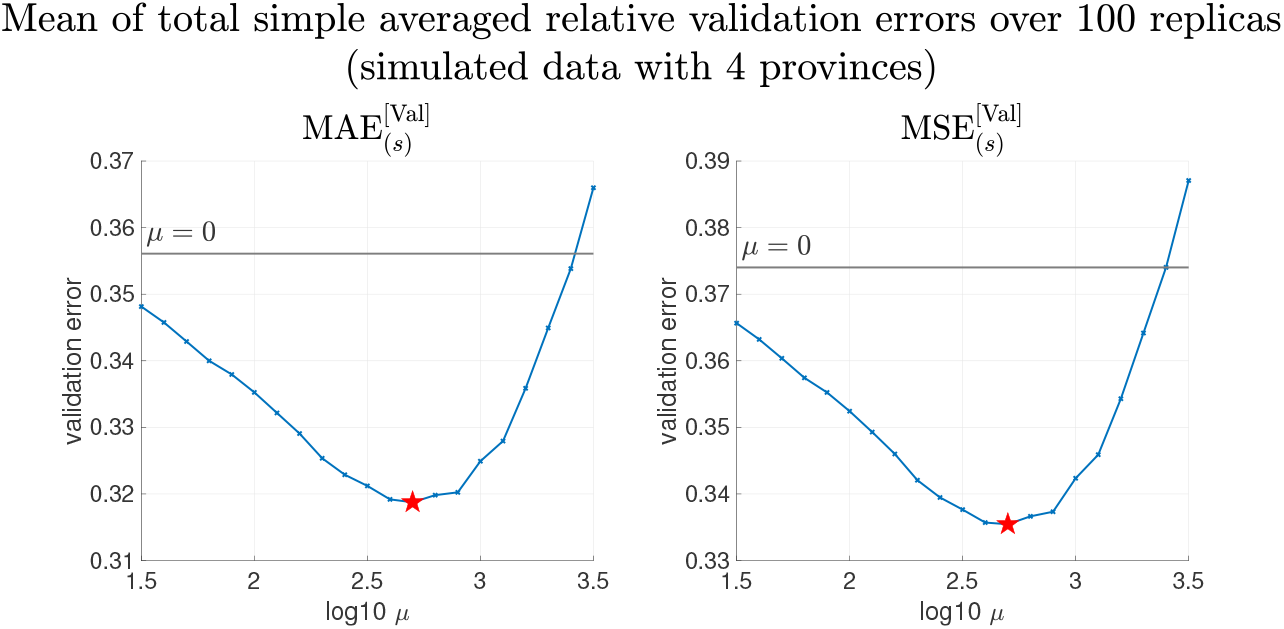
Mean of simply averaged validation errors 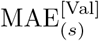 and 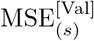 against *µ* (computed by (A5)) over 100 replicas for simulated data (four provinces). The horizontal lines show the values of the mean of the validation errors when *µ* = 0.

**Figure A2:**
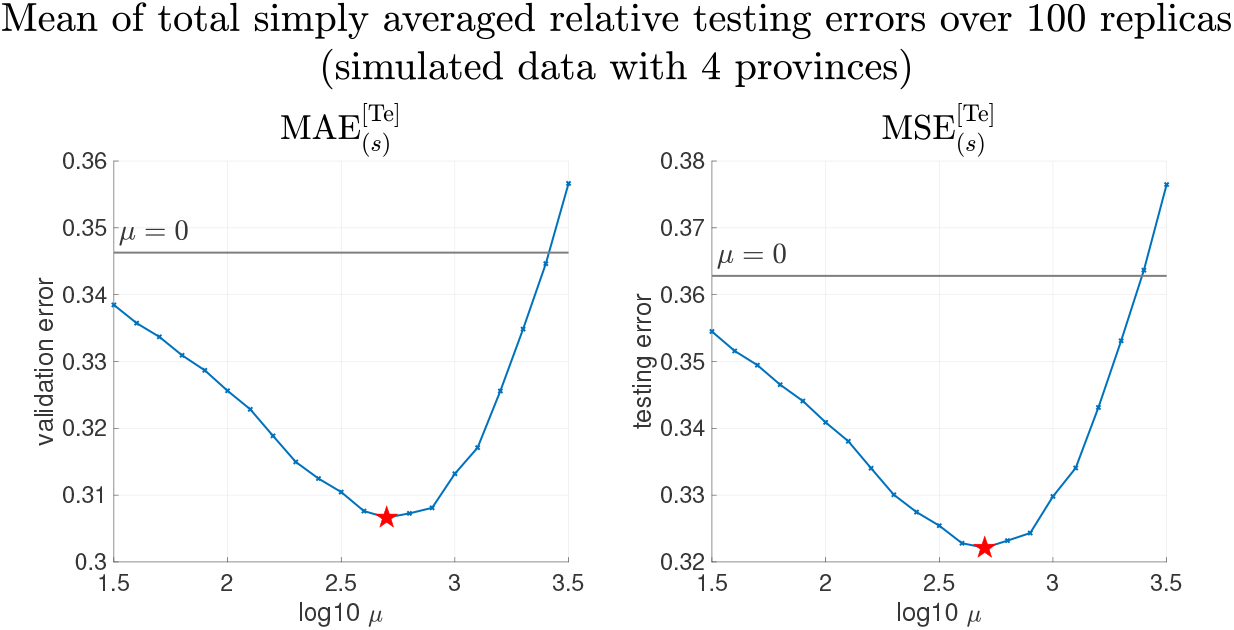
Mean of simply averaged testing errors 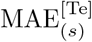 and 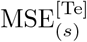 against *µ* (computed by (A5)) over 100 replicas for simulated data (four provinces). The horizontal lines show the values of the mean of the testing errors when *µ* = 0.

**Figure A3:**
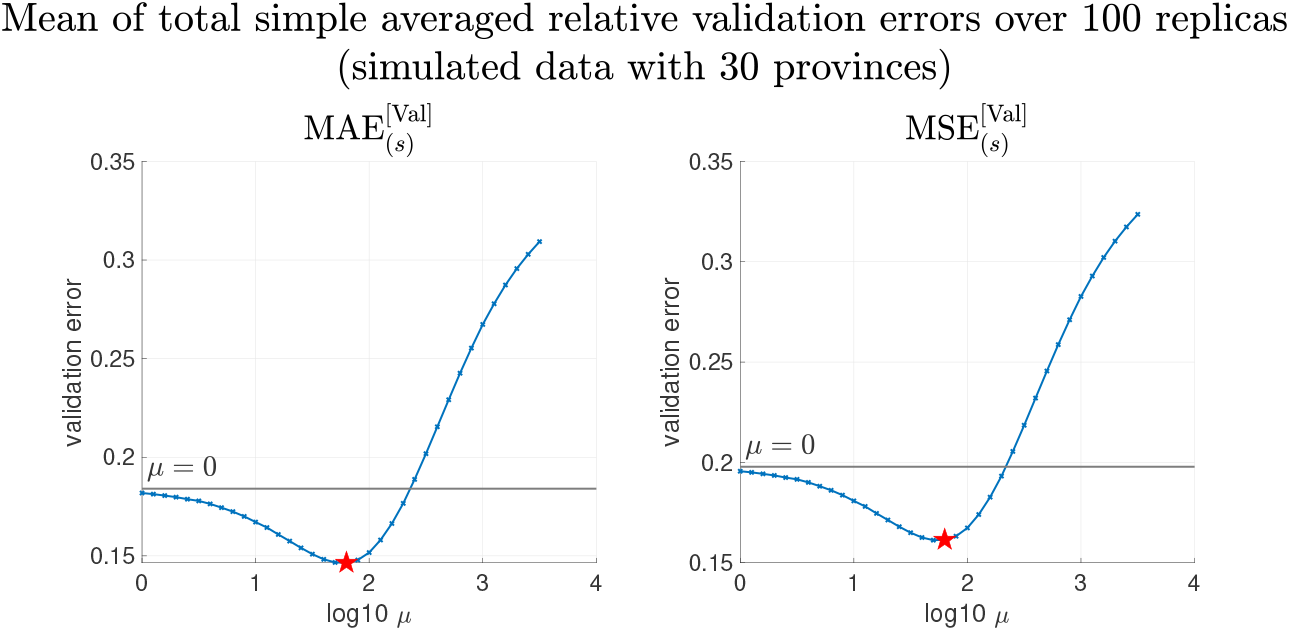
Mean of simply averaged validation errors 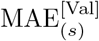 and 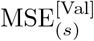 against *µ* (computed by (A5)) over 100 replicas for simulated data (thirty provinces). The horizontal lines show the values of the mean of the validation errors when *µ* = 0.

**Figure A4:**
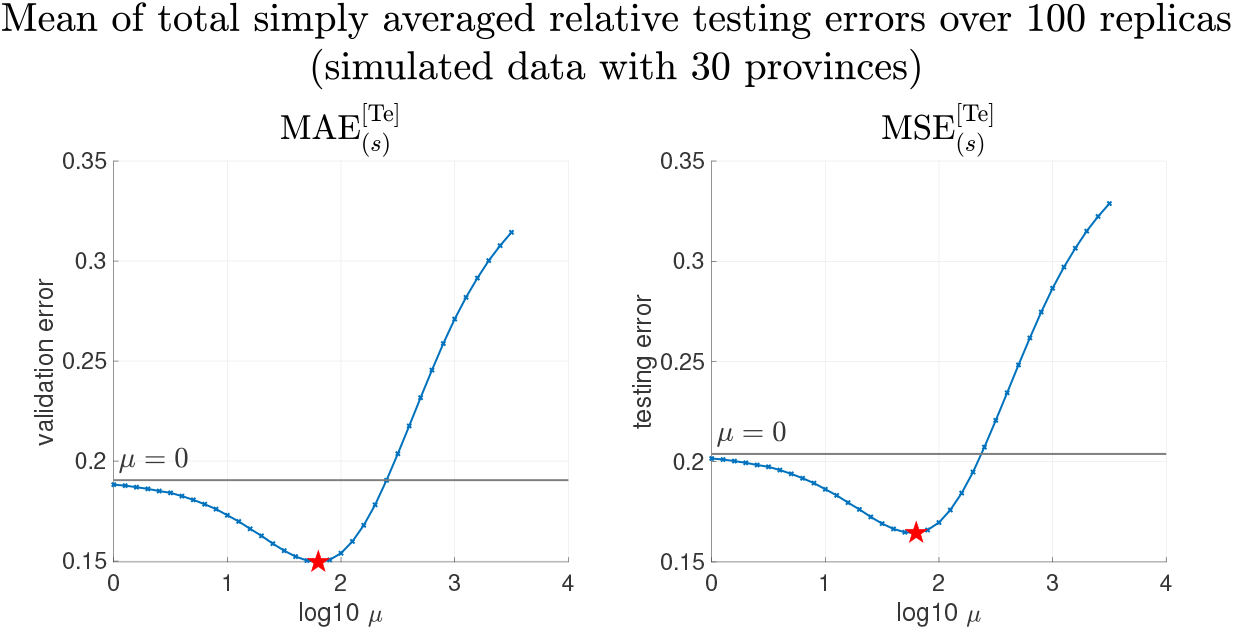
Mean of simply averaged testing errors 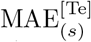 and 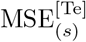 against *µ* (computed by (A5)) over 100 replicas for simulated data (thirty provinces). The horizontal lines show the values of the mean of the testing errors when *µ* = 0.

**Figure A5:**
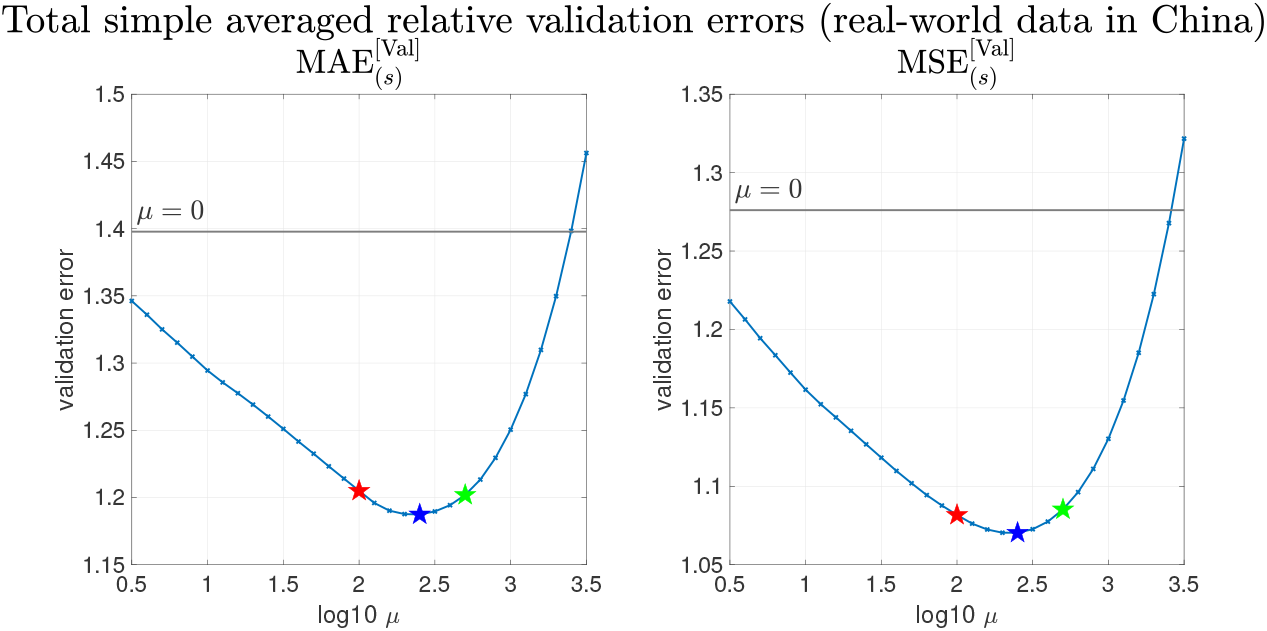
Simply averaged validation errors 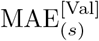 and 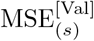 against *µ* (computed by (A5)) for real-world data in China. The horizontal lines show the values of the validation errors when *µ* = 0.

**Figure A6:**
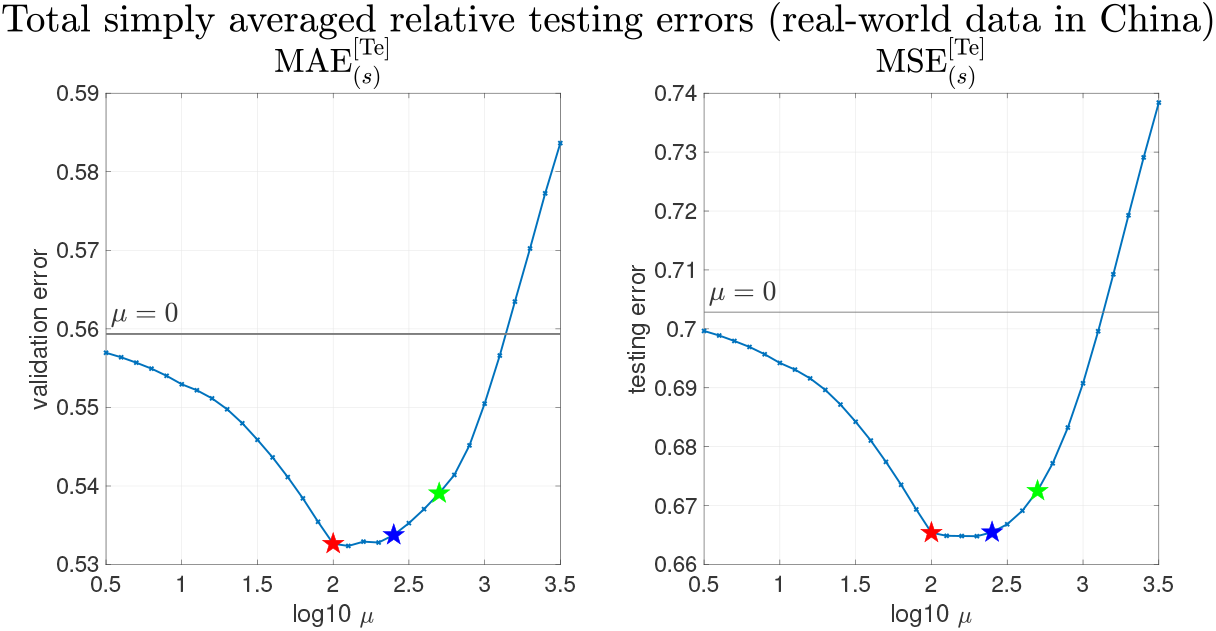
Simply averaged testing errors 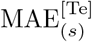 and 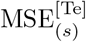 against *µ* (computed by (A5)) for real-world data in China. The horizontal lines show the values of the testing errors when *µ* = 0.

**Figure A7:**
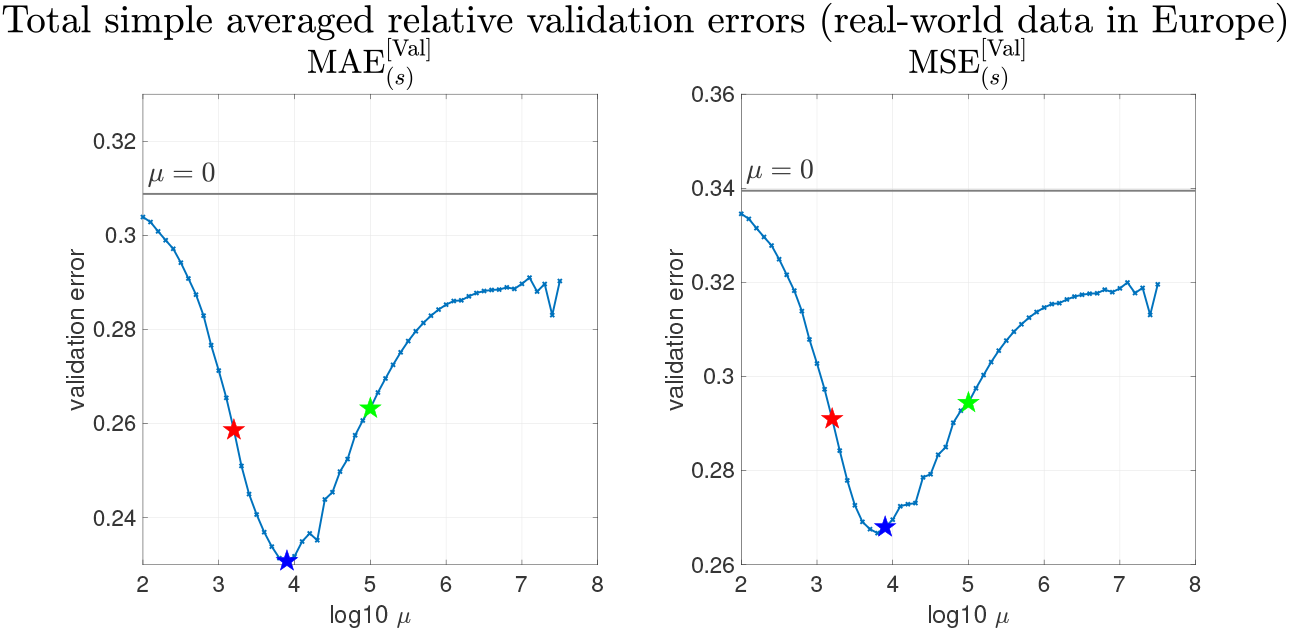
Simply averaged validation errors 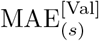 and 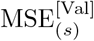 against *µ* (computed by (A5)) for real-world data in Europe. The horizontal lines show the values of the validation errors when *µ* = 0.

**Figure A8:**
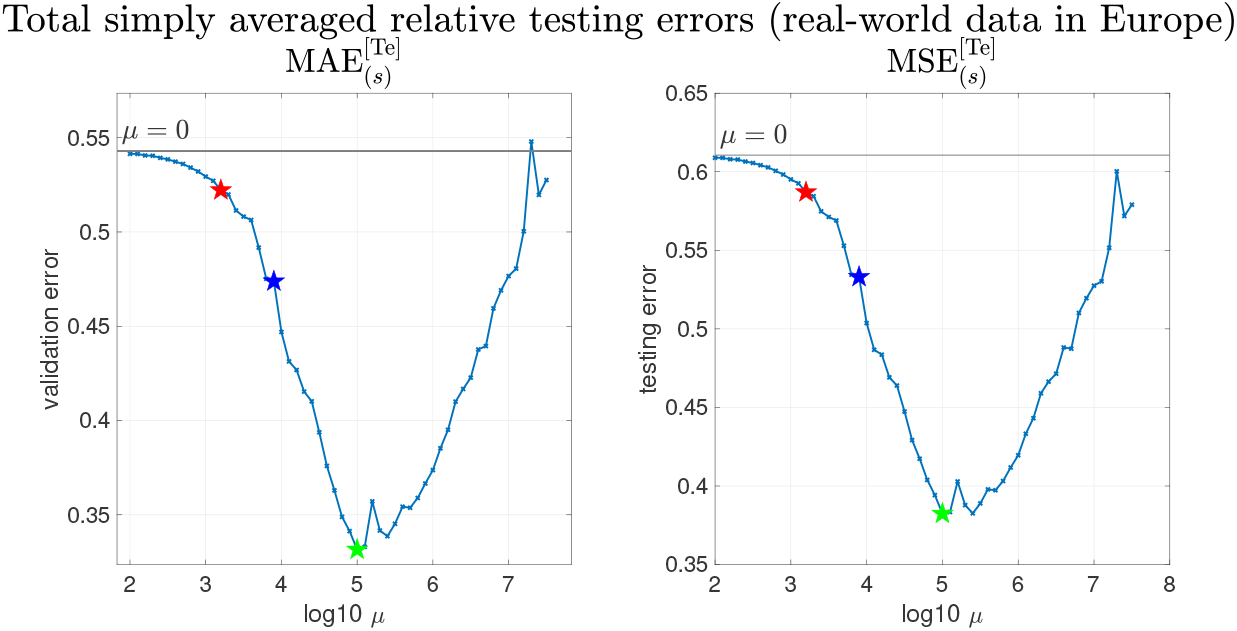
Simply averaged testing errors 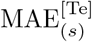 and 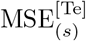 against *µ* (computed by (A5)) for real-world data in Europe. The horizontal lines show the values of the testing errors when *µ* = 0.

## References

[1] Balcan, D. et al. Modeling the spatial spread of infectious diseases: The global epidemic and mobility computational model. Journal of computational science 1, 132–145 (2010).

[2] World Health Organization (WHO). https://covid19.who.int.

[3] Keeling, M. J. & Rohani, P. Modeling infectious diseases in humans and animals (Princeton university press, 2011).

[4] Gomes, M. F. et al. Assessing the international spreading risk associated with the 2014 west african ebola outbreak. PLoS currents 6 (2014).

[5] Pastore-Piontti, A. et al. Real-time assessment of the international spreading risk associated with the 2014 west african ebola outbreak. In Mathematical and Statistical Modeling for Emerging and Re-emerging Infectious Diseases, 39–56 (Springer, 2016).

[6] Chinazzi, M. et al. The effect of travel restrictions on the spread of the 2019 novel coronavirus (covid-19) outbreak. Science 368, 395–400 (2020).

[7] Colizza, V. et al. Estimate of novel influenza a/h1n1 cases in mexico at the early stage of the pandemic with a spatially structured epidemic model. PLoS currents 1, RRN1129–RRN1129 (2009).

[8] Balcan, D. et al. Modeling the critical care demand and antibiotics resources needed during the fall 2009 wave of influenza a(h1n1) pandemic. PLoS currents 1, RRN1133–RRN1133 (2009).

[9] Bajardi, P. et al. Modeling vaccination campaigns and the fall/winter 2009 activity of the new a(h1n1) influenza in the northern hemisphere. Emerging health threats journal 2, e11–e11 (2009;2008;2010;).

[10] Tizzoni, M. et al. Real-time numerical forecast of global epidemic spreading: Case study of 2009 a/h1n1pdm. BMC medicine 10, 165–165 (2012).

[11] Poletto, C. et al. Assessment of the middle east respiratory syndrome coronavirus (mers-cov) epidemic in the middle east and risk of international spread using a novel maximum likelihood analysis approach. Eurosurveillance 19, 20824 (2014).

[12] Balcan, D. et al. Seasonal transmission potential and activity peaks of the new influenza a (h1n1): a monte carlo likelihood analysis based on human mobility. BMC medicine 7, 1–12 (2009).

[13] Dong, E., Du, H. & Gardner, L. An interactive web-based dashboard to track covid-19 in real time. The Lancet infectious diseases 20, 533–534 (2020).

[14] Zhang, Q. et al. Forecasting seasonal influenza fusing digital indicators and a mechanistic disease model. In Proceedings of the 26th international conference on world wide web, 311–319 (2017).

[15] Vespignani, A. Predicting the behavior of techno-social systems. Science 325, 425–428 (2009).

[16] Donnat, C. & Holmes, S. Modeling the heterogeneity in covid-19’s reproductive number and its impact on predictive scenarios. Journal of Applied Statistics 1–29 (2021).

[17] Tkachenko, A. V. et al. Time-dependent heterogeneity leads to transient suppression of the covid-19 epidemic, not herd immunity. Proceedings of the National Academy of Sciences 118 (2021).

[18] Moreno, Y., Pastor-Satorras, R. & Vespignani, A. Epidemic outbreaks in complex heterogeneous networks. The European Physical Journal B-Condensed Matter and Complex Systems 26, 521–529 (2002).

[19] Kermack, W. O. & McKendrick, A. G. A contribution to the mathematical theory of epidemics. Proceedings of the royal society of london. Series A, Containing papers of a mathematical and physical character 115, 700–721 (1927).

[20] Boschi, T., Di Iorio, J., Testa, L., Cremona, M. A. & Chiaromonte, F. Functional data analysis characterizes the shapes of the first covid-19 epidemic wave in italy. Scientific reports 11, 1–15 (2021).

[21] Carroll, C. et al. Time dynamics of covid-19. Scientific reports 10, 1–14 (2020).

[22] Cremona, M. A. & Chiaromonte, F. Probabilistic k-mean with local alignment for clustering and motif discovery in functional data. arXiv preprint 1808.04773 (2018).

[23] Hilton, J. & Keeling, M. J. Estimation of country-level basic reproductive ratios for novel coronavirus (sars-cov-2/covid-19) using synthetic contact matrices. PLoS computational biology 16, e1008031 (2020).

[24] Szapudi, I. Heterogeneity in sir epidemics modeling: superspreaders and herd immunity. Applied Network Science 5, 1–12 (2020).

[25] Volpert, V., Banerjee, M. & Sharma, S. Epidemic progression and vaccination in a heterogeneous population. application to the covid-19 epidemic. Ecological Complexity 100940 (2021).

[26] Hou, X. et al. Intracounty modeling of covid-19 infection with human mobility: Assessing spatial heterogeneity with business traffic, age, and race. Proceedings of the National Academy of Sciences 118 (2021).

[27] Chen, S., Li, Q., Gao, S., Kang, Y. & Shi, X. State-specific projection of covid-19 infection in the united states and evaluation of three major control measures. Scientific reports 10, 1–9 (2020).

[28] National Health Commission of the People’s Republic of China. http://en.nhc.gov.cn/antivirusfight.html.

[29] Chinese Center for Disease Control and Prevention. https://weekly.chinacdc.cn/news/TrackingtheEpidemic.htm.

[30] European Centre for Disease Prevention and Control. https://www.ecdc.europa.eu/en/geographical-distribution-2019-ncov-cases.

[31] Tian, H. et al. An investigation of transmission control measures during the first 50 days of the covid-19 epidemic in china. Science 368, 638–642 (2020).

[32] Kraemer, M. U. et al. The effect of human mobility and control measures on the covid-19 epidemic in china. Science 368, 493–497 (2020).

[33] World Health Organization (WHO). Enhancing response to omicron sars-cov-2 variant. https://www.who.int/publications/m/item/enhancing-readiness-for-omicron-(b.1.1.529)-technical-brief-and-priority-actions-for-member-states.

[34] Zhang, Y. et al. Prediction of the covid-19 outbreak based on a realistic stochastic model. medRxiv (2020).

[35] Kurtz, T. G. Solutions of ordinary differential equations as limits of pure jump markov processes. Journal of applied Probability 7, 49–58 (1970).

[36] Kurtz, T. G. Limit theorems for sequences of jump markov processes approximating ordinary differential processes. Journal of Applied Probability 8, 344–356 (1971).

[37] He, S., Peng, Y. & Sun, K. Seir modeling of the covid-19 and its dynamics. Nonlinear dynamics 101, 1667–1680 (2020).

[38] López, L. & Rodo, X. A modified seir model to predict the covid-19 outbreak in spain and italy: simulating control scenarios and multi-scale epidemics. Results in Physics 21, 103746 (2021).

[39] Yang, Z. et al. Modified seir and ai prediction of the epidemics trend of covid-19 in china under public health interventions. Journal of thoracic disease 12, 165 (2020).

[40] He, X. et al. Temporal dynamics in viral shedding and transmissibility of covid-19. Nature medicine 26, 672–675 (2020).

[41] National Bureau of Statistics of China. Annual data by province (2019). http://www.stats.gov.cn/english/Statisticaldata/AnnualData/.

[42] Baidu Qianxi. https://qianxi.baidu.com/#/2020chunyun.

[43] Johns Hopkins University. Covid-19 data repository by the center for systems science and engineering (csse) at johns hopkins university (2020). https://github.com/CSSEGISandData/COVID-19.

[44] Wikipedia. List of european countries by population. https://en.wikipedia.org/wiki/List_of_European_countries_by_population.

[45] Information Office of Hubei Provincial People’s Government. Prevention and control of pneumonia outbreak of new coronary virus infection (2020). https://www.hubei.gov.cn/hbfb/xwfbh/202002/t20200210_2023490.shtml.

[46] Blavatnik School of Government. Covid-19 government response tracker. https://www.bsg.ox.ac.uk/research/research-projects/covid-19-government-response-tracker.

[47] Kendall, D. G. Deterministic and stochastic epidemics in closed populations. In Proceedings of the Third Berkeley Symposium on Mathematical Statistics and Probability, Volume 4: Contributions to Biology and Problems of Health, 149–165 (University of California Press, 1956).

[48] Bailey, N. T. A simple stochastic epidemic. Biometrika 193–202 (1950).

[49] Bartlett, M. Some evolutionary stochastic processes. Journal of the Royal Statistical Society. Series B (Methodological) 11, 211–229 (1949).

[50] Britton, T. et al. Stochastic epidemic models with inference (Springer, 2019).

